# Predicting interstitial lung disease progression in patients with systemic sclerosis using attentive neural processes - a EUSTAR study

**DOI:** 10.1101/2024.04.25.24306365

**Authors:** Ahmed Allam, Aron N. Horvath, Matthias Dittberner, Cécile Trottet, Elise Siegert, Vincent Sobanski, Patricia Carreira Delgado, Dagna Lorenzo, Vanessa Smith, Serena Guiducci, Nicolas Hunzelmann, Anna-Maria Hoffmann-Vold, Simona Trugli, Ana Maria Gheorghiu, Agachi Svetlana, Camillo Ribi, Michael Krauthammer, Britta Maurer, EUSTAR Collaborators

## Abstract

**Background:** Systemic sclerosis (SSc) is an autoimmune disease with high mortality with lung involvement being the primary cause of death. Progressive interstitial lung disease (ILD) leads to a decline in lung function (forced vital capacity, FVC% predicted) with risk of respiratory failure. These patients could benefit from an early and tailored pharmacological intervention. However, up to date, tools for prediction of individual FVC changes are lacking. In this paper, we aimed at developing a trustworthy machine learning system that is able to guide SSc management by providing not only robust FVC predictions, but also uncertainty quantification (i.e. the degree of certainty of model prediction) as well as similarity-based explainability for any patient P (i.e. a list of past SSc patients with similar FVC trajectories like P). We further aimed to identify the key clinical factors influencing the model’s predictions and to use model-guided data representation to identify SSc patients with similar sequential FVC measurements.

**Methods:** We trained and evaluated machine learning (ML) models to predict SSc-ILD trajectory as measured by FVC% predicted values using the international SSc database managed by the European Scleroderma Trials and Research group (EUSTAR), which comprises clinical, laboratory and functional parameters. EUSTAR records patients’ data in annual assessment visits, and, given any visit, we aimed at predicting the FVC value of a patient’s subsequent visit, taking into account all available patient data (i.e. baseline and follow-up visit data up to the time point where we make the prediction). For training of our ML models, we included 2220 SSc patients that had at least 3 recorded visits in the EUSTAR database, were at least 18 years old, had confirmed ILD and sufficient clinical documentation. We developed sequential ML models implementing the attentive neural process formalism with either a recurrent (ANP RNN) or transformer encoder (ANP transformer) architecture. We compared these architectures with baseline sequential models including gated recurrent neural networks (RNNs) and multi-head self-attention transformer-based networks. Baseline non-sequential models included tree-based models such as gradient boosting trees, and regression-based models with varying regularization schemes. Our experiments used stratified 5-fold cross-validation to train and test the models using the average root mean squared error (RMSE), weighted RMSE, and mean absolute error (MAE) as performance metrics. We computed the coverage and Winkler score for uncertainty quantification, SHAP values for grading the input features importance and used the data embeddings of the ANP architectures for both similarity-based explainability and the identification of similar SSc patient journeys.

**Results:** Patients’ baseline FVC scores ranged from 22 to 150% predicted with a mean (SD) of 90.53% predicted (21.52). Our deep learning models showed better performance for FVC forecasting, compared to tree- and regression-based models. The top performing ANP RNN architecture was able to closely model future FVC values with average (SD) performance of 8.240 (0.168) weighted RMSE and 6.94 (0.190) MAE that was further used as feature generator for a logistic regression trained to predict a FVC% decline of at least 10% points achieving 0.704 AUC score. In comparison, a naïve baseline using the mean FVC value as a predictor achieved much lower FVC forecasting capabilities, with 18.718 (0.317) weighted RMSE, and 17.619 (0.599) MAE. SHAP value analysis indicated that prior FVC measurements, diffusion of carbon monoxide (DLCO) values, skin involvement, age, anti-centromere positivity, dyspnea and CRP-elevation contributed most to deep-learning-based FVC predictions. Regarding uncertainty quantification, ANP RNN achieved 79% coverage (i.e. the model would provide uncertainty estimates that included the true future FVC value in 79 out of 100 predictions) out of the box, and 90% using an additional conformal prediction module with an corresponding Winkler score of 892 (indicating the width of the uncertainty estimate plus penalty for mistakes), smaller than any other model at the same coverage level. We further demonstrate how the data abstraction provided by the ANP RNN model (embeddings) allows for deriving similar patient trajectories (for similarity-based explanation).

**Conclusions:** Our study demonstrates the feasibility of FVC forecasting and thus the ability to predict ILD trajectories in individual SSc patients using deep learning. We show that model predictions can be paired with uncertainty quantification and similarity-based model explainability, which are crucial elements for deploying trustworthy ML algorithms. Our study is thus an important first step towards reliable automated ILD trajectory (i.e. FVC%) prediction system with potential clinical utility.

## 1 Introduction

Systematic Sclerosis (SSc) is a chronic autoimmune disease with prominent characteristics of vascular damage, dysregulation of the immune system and progressive fibrosis affecting different tissues and organs [1, 2]. One of the consequences of the excessive deposition of extracellular matrix is the development of interstitial lung disease (ILD) leading to progressive structural and functional worsening of the affected tissue. With ILD as one of the major complications and leading causes of death of SSc [3], patients diagnosed with SSc are subjected to repeated comprehensive clinical assessments to identify early signs of deterioration of lung function or to monitor disease progression in order to enable optimized treatment and course of actions. The current practice includes high resolution computer tomography (HRCT) of the chest to diagnose and asses the extent of lung fibrosis in combination with pulmonary function tests (PFTs) to assess disease severity. Recently, computational analysis of HRCT-derived metadata, i.e. radiomics, showed prognostic potential for the prediction of progression-free survival of patients with SSc-ILD [4]. The prognostic value of PFTs have been evaluated by N. Goh et al, who concluded that changes in forced vital capacity (FVC) and diffusing capacity of carbon monoxide (DLCO) compared to the baseline can be used for the assessment of ILD progression [5], [6]. Since there is a large variation between patients with respect of disease severity and progression (progressive ILD associated with higher mortality rate as compared with stable disease course [7][8]), the key of successful disease management depends on the ability to assess and predict the lung function trajectory of the individual SSc-ILD patient in order to provide optimized management strategies.

Recent studies reported a few biomarkers including anti-topoisomerase I and multiple inflammatory markers as predictor candidates for SSc-ILD progression, however their further evaluation is still required [9, 10]. Wu et al. developed the SPAR prediction model deriving SpO2 after 6 minutes walk and presence of arthritis as two independent factors for SSc-ILD progression [11]. Furthermore, Kaenmuang et al. have identified gender (male) and no previous aspirin treatment in a relative small unique cohort (78 patients) as alternative predictive factors [12]. On the other hand, Hoffman-Vold et al., separately examined risk factors predictive ILD progression within 1 year and 5 years [13] using a significantly larger cohort from the EUSTAR database (826 ILD patients). Their analysis concluded that presence of reflux/dysphagia, high baseline of mRSS and value of FVC at 12 months as well as male sex, older age and higher DLCO were significant predictive factors of FVC decline within the 5 year time window. However, despite these significant efforts, to date there is no confirmed clinical parameter or biomarker that could predict SSc-ILD progression on an individual level as well as there is no clear consensus on screening frequency and methodology [9].

The underlying reasons include not only the different size and characteristics of cohorts used in the aforementioned retrospective studies [12, 13, 11], but potentially the used data analysis approaches which are mainly based on uni- and multivariate linear/logistic regression. In recent years, machine learning (ML) has shown great potential to extract actionable insights from medical data which can be used to support clinical decision making [14, 15]. For example, Yan et al. successfully combined genetic and imaging data for predicting of progression of age-related macular degeneration [16]. Furthermore, ML frameworks were developed to predict rapid coronary plaque progression to identify patients at risk, as well as to assess the importance of clinical parameters [17]. Similarly, Garaiman et al. assessed the performance of vision transformer (deep learning image based model) for identifying distinct signs of microangiopathy using nailfold capilloroscopy images of patients with SSc [18].

### 1.1 Our contribution

In this study, we modeled SSc-ILD trajectories as a regression problem by developing ML algorithms for the prediction of future FVC% values. We built patients’ trajectories (i.e. timelines) out of the recorded events extracted from the patient’s information while preserving the temporal progression and the history for every patient in the database. As part of our contribution, (1) we propose an adaptation for the attentive neural process formulation [19] to model the FVC% predicted values in patients’ trajectories. (2) We further compare our proposed model architecture to a wide array of ML models such as gated recurrent neural network [20, 21], transformer network [22] and other baseline ML models. (3) We then study and assess the uncertainty estimates of the predicted outcomes by our model and contrast it to a common post-hoc approach for uncertainty estimation used in neural network models. (4) We further experiment with conformal prediction procedure [23, 24] to improve the uncertainty estimates provided by the trained models, and (5) lastly report on the main features contributing in model’s prediction decision using (a) SHAP values and (b) data embedding computed by our model for similarity-based explainability.

## 2 Materials and Methods

### 2.1 Data source and data extraction

Our study used the EUSTAR database that has been extensively described previously in [25, 26]. Briefly, the database is managed and maintained by the European Scleroderma Trials and Research group and contains data from more than 15’000 patients from more than 200 sites. The database provides longitudinal observational data documenting each patient’s visit including sociodemographics, clinical, laboratory and functional data, and information on therapy.

In our study, we included all patients who fulfilled the following criteria: (1) aged ≥ 18 years; (2) fulfilled criteria of the 2013 American College of Rheumatology/European League Against Rheumatism SSc classification [27] (3) presence of ILD confirmed by high resolution HRCT (high resolution computed tomography) or X-ray of the chest; (4) documented FVC% predicted and DLCO% predicted measurements; (5) availability of at least 3 visits in their timeline.

#### 2.1.1 Patients trajectory processing

The extracted patient information was preprocessed to build trajectories (i.e. timelines) out of the recorded events while preserving the temporal progression and the history for every patient. Formally, we denote each visit at time *t* by a feature vector 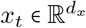 encoding sociodemographics, clinical data, and information on medication used. For each of these visits in the trajectory, the aim was to predict the future FVC% value recorded in the subsequent visit at *t* + *δt* denoted by 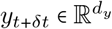 .

### 2.2 Patients’ characteristics

The final dataset included 2220 patients with an average age of 53.42 years (at baseline) with 83% females. The demographic characteristics of patients included: age, sex, smoking habits and ethnicity that were simplified as reported in Table 1. Patients’ characteristics assessed at every visit including the FVC% and DLCO% measurements are summarized in Tables 2 and 3. On average, the enrolled patients had 5.94 ± 2.96 visits, ranging from 3 to 19 visits as reported in Figure 1. A plot of the whole patient trajectories is reported in Figure 2 and Figure 11 in Appendix. Additionally, presence or absence of a selected set of treatment data was included in the modeling. A list of all features used to train the ML models is reported in Appendix E.

**Table 1:** Demographic characteristics of all patients.

| Parameter | Total (N=2220) | Missing data (%) |
| --- | --- | --- |
| Age*, years (SD) | 53.42 (12.91) | 0 |
| Male, n (%) | 370 (16.67) | 0 |
| Visits, n (SD) | 5.92 (2.97) | 0 |
| Smoker, yes (no) | 603 (1418) | 8.96 |
| Ethnicity, n |  | 10.95 |
| White | 1880 |  |
| Hispanic | 12 |  |
| Asian | 36 |  |
| Black | 29 |  |
| Middle-eastern | 7 |  |
| Maghrebis | 15 |  |
| Other | 12 |  |

**Table 2:** Clinical data - numerical features.

| Parameter | Mean | Std | Missing values (%) |
| --- | --- | --- | --- |
| Erythrocyte sedimentation rate (mm/h) | 23.32 | 18.29 | 13.20 |
| FVC% predicted | 90.53 | 21.52 | 0.00 |
| DLCO% predicted | 62.32 | 19.61 | 0.00 |

**Table 3:** Clinical data - categorical features.

| Parameter | Categories | Values (%) |
| --- | --- | --- |
| Raynaud's present | No | 5.25 |
| Raynaud's present | Yes | 92.03 |
| Raynaud's present | Missing | 2.72 |
| Esophageal symptoms (dysphagia, reflux) | No | 39.72 |
| Esophageal symptoms (dysphagia, reflux) | Yes | 59.22 |
| Esophageal symptoms (dysphagia, reflux) | Missing | 1.06 |
| ANA positive | No | 2.72 |
| ANA positive | Yes | 87.92 |
| ANA positive | Missing | 9.36 |
| RNA Polymerase III positive | No | 52.78 |
| RNA Polymerase III positive | Yes | 3.13 |
| RNA Polymerase III positive | Missing | 44.09 |
| Muscle weakness | No | 80.8 |
| Muscle weakness | Yes | 15.6 |
| Muscle weakness | Missing | 3.6 |
| Dyspnea (significant) | No | 82.86 |
| Dyspnea (significant) | Yes | 11.33 |
| Dyspnea (significant) | Missing | 5.81 |
| CRP-Elevation | No | 69.71 |
| CRP-Elevation | Yes | 21.83 |
| CRP-Elevation | Missing | 8.46 |
| Renal crisis | No | 96.9 |
| Renal crisis | Yes | 1.12 |
| Renal crisis | Missing | 1.97 |
| Joint synovitis | No | 87.7 |
| Joint synovitis | Yes | 9.75 |
| Joint synovitis | Missing | 2.55 |
| Tendon friction rubs | No | 88.77 |
| Tendon friction rubs | Yes | 6.93 |
| Tendon friction rubs | Missing | 4.3 |
| Extent of skin involvement | Only_sclerodactyly | 9.17 |
| Extent of skin involvement | Limited_cutaneous_involvement | 44.26 |
| Extent of skin involvement | Diffuse_cutaneous_involvement | 35.76 |
| Extent of skin involvement | No_skin_involvement | 7.04 |
| Extent of skin involvement | Missing | 3.77 |
| Digital Ulcers | Previously | 23.14 |
| Digital Ulcers | Current | 7.94 |
| Digital Ulcers | Never | 24.84 |
| Digital Ulcers | Missing | 44.08 |
| Dyspnea (NYHA-stage) | 1 | 45.66 |
| Dyspnea (NYHA-stage) | 2 | 37.52 |
| Dyspnea (NYHA-stage) | 3 | 10.07 |
| Dyspnea (NYHA-stage) | 4 | 1.26 |
| Dyspnea (NYHA-stage) | Missing | 5.49 |

**Figure 1:**
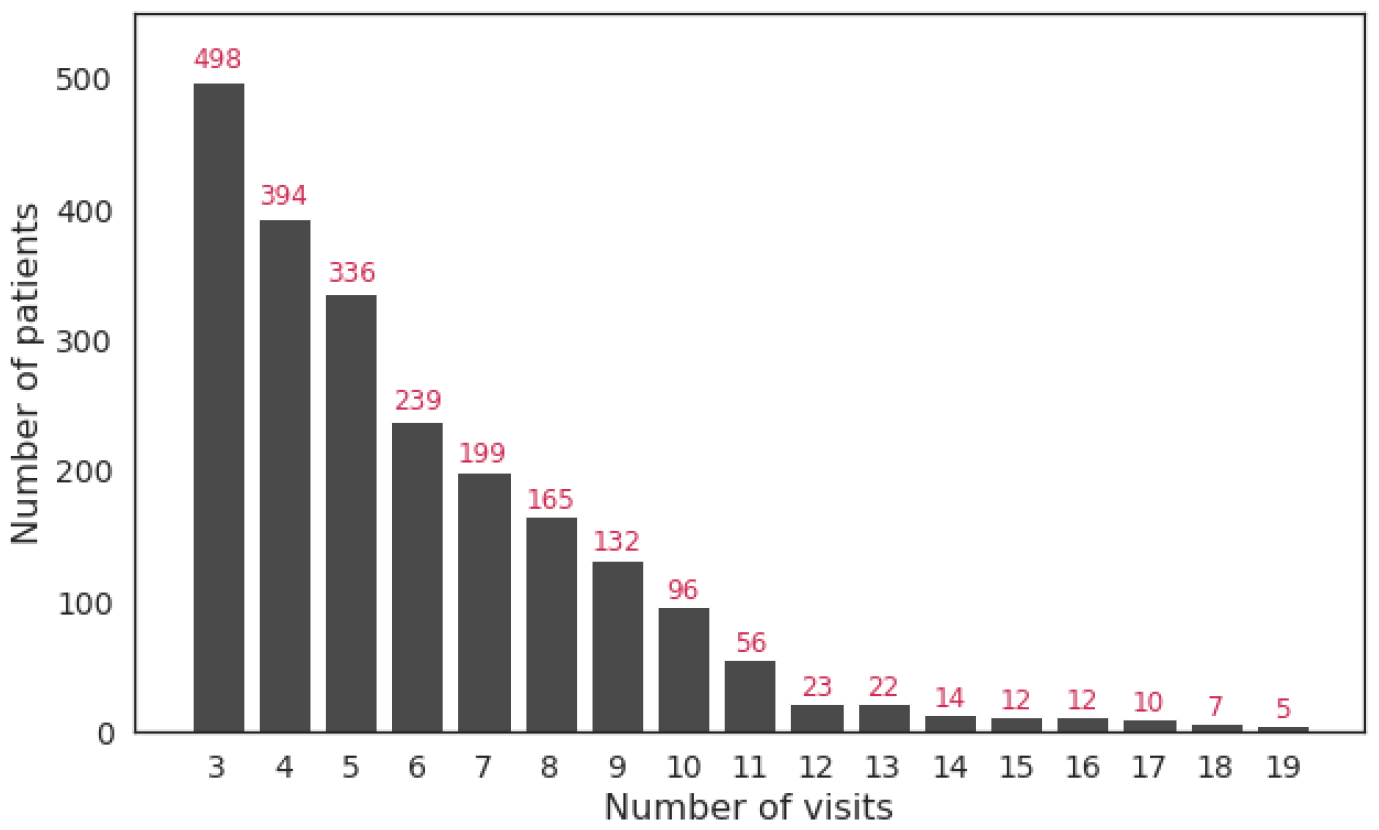
Distribution of the number of visits across patients.

**Figure 2:**
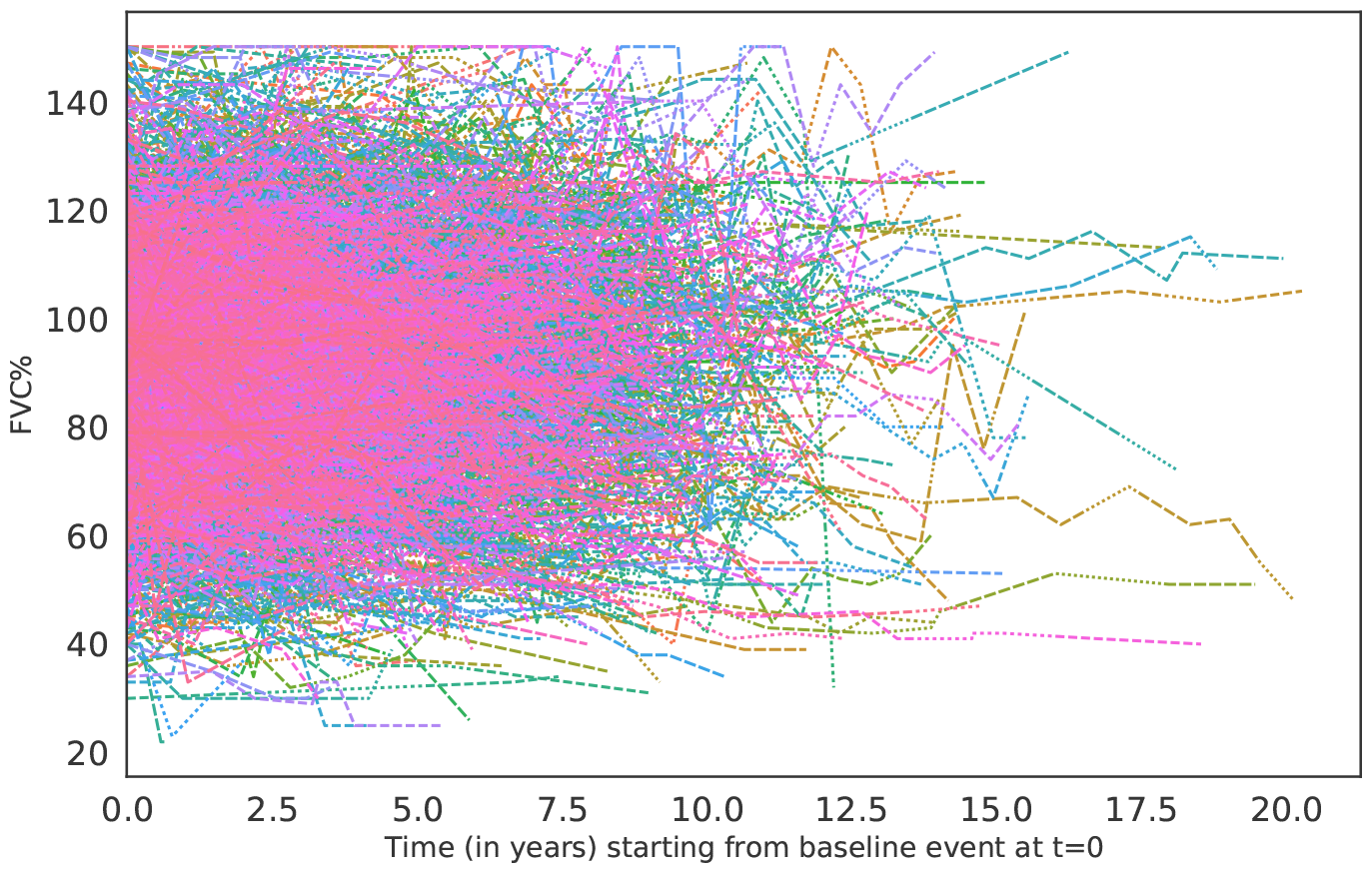
Line plot of patients’ trajectories spanning 20 years of events.

### 2.3 Disease progression modelling

Given a patient’s temporally ordered sequence of visits 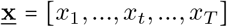 where each visit is represented by a *d*_*x*_-dimensional feature vector 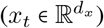, we aim at predicting the next visit’s recorded outcome *y*_*T* +*δT*_ (i.e. FVC% predicted value) based on all visits available in the past trajectory (in this case *t* ∈ [1, ⋯, *T*]).

Hence, for a given visit *x*_*t*_, a model will use all available visits up to *t* (i.e. [*x*_1_, ⋯, *x*_*t*_]) to predict the subsequent visit outcome at *t* + *δt* denoted by 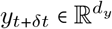 . As a result, an input sequence represented by matrix 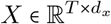 will generate an outcome sequence matrix 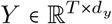. Given a training set 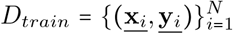 consisting of *N* input-output sequence pairs, the goal is to learn a model (i.e. function map *f*) by minimizing an objective function *L* (*f, D*_*train*_) that measures the discrepancy between sequence of reference outcome values <u>**y**_*i*_</u> and its corresponding predicted outcome values <u>**ŷ**_*i*_</u> in the training dataset. In the following sections, we describe the different modelling approaches used in this study to learn a parametrized function *f* (***θ***) minimizing the defined differentiable objective function by finding “optimal weights” ***θ*** where ***θ*** = arg min_***θ***_ *L*(*f, D*_*train*_).

### 2.4 Recurrent neural network (RNN)

We used recurrent neural networks (RNN) that is suited for modeling sequential and temporal data with varying length [28, 29]. RNNs computes a hidden vector at each time step (i.e. state vector *h*_*t*_ at time *t*), representing a history or context summary of the sequence using the input and hidden states vector form the previous time step. This allows the model to learn long-range dependencies where the network is unfolded as many times as the length of the sequence it is modeling. In this work, we used gated recurrent unit (GRU) [20, 30] to overcome the vanishing/exploding gradient challenges [31, 32, 29] by updating the computation mechanism of the hidden state vector *h*_*t*_ through the specified equations in Appendix C. An output layer is added on top that maps the hidden state vector representation to the outcome representing FVC value at future time point. We will refer to the GRU based model by RNN throughout the paper.

### 2.5 Transformer network

Another model architecture we explored in modelling disease progression is Transformer network [22]. The model has three main blocks: An (1) **Embedding block** that embeds both the *features* and corresponding *absolute position* to a dense vector representation (we also experimented with *time embedding* variation replacing position embedding component). An (2) **Encoder block** that contains (a) a multi-head self-attention layer, (b) layer normalization & residual connections, and (c) feed-forward network. Lastly, an (3) **Output block** representing a regression layer for predicting the subsequent visits FVC% predicted value. A formal description of each component of the model is described in their respective sections in the Appendix C.

### 2.6 Attentive Neural Processes (ANP)

Attentive Neural Processes (ANP) [19] is an extension to Neural Processes [33], an approach that learns a distribution over functions mapping the input to output from a training set (i.e. learning a posterior distribution over *f* the underlying function mapping input to output) that is further used to make inference for test points. ANP defines an infinite family of conditional distributions conditioning on arbitrary number of *contexts* (i.e. set of input-output pairs (**x**_*C*_, **y**_*C*_) = {(*x*_1_, *y*_1_), (*x*_2_, *y*_2_), ⋯, (*x*_*C*_, *y*_*C*_)}) to model arbitrary number of *targets* (**x**_*M*_, **y**_*M*_) ={( *x*_1_, *y*_1_), { (*x*_2_, *y*_2_), ⋯, (*x*_*C*_, *y*_*C*_), ⋯, (*x*_*M*_, *y*_*M*_) } invariant to the ordering of both the contexts and targets where *C* ⊂ *M* Eq. 25 in Appendix C. In this work, we adapt ANP to model patient trajectories (i.e. timeseries data) where causal temporal ordering is preserved and we describe the adaptation of the modeling approach from this perspective.

ANP comprises of an (1) **Encoder block** that uses two paths (a) *deterministic* and (b) *latent path*, and (2) **Decoder block** that maps the computed representation from the encoder block to the the target output (Figure 3). In this study, we first used a time series encoder that embeds the raw input for both the context and target events, then pass the learned representations to the encoder blocks Φ and Ω (as shown in Figure 3). We experimented with two model variations: the first used a gated RNN for all the encoder blocks and is denoted by ANP RNN, and the second used transformer based encoder blocks and is denoted by ANP transformer.

**Figure 3:**
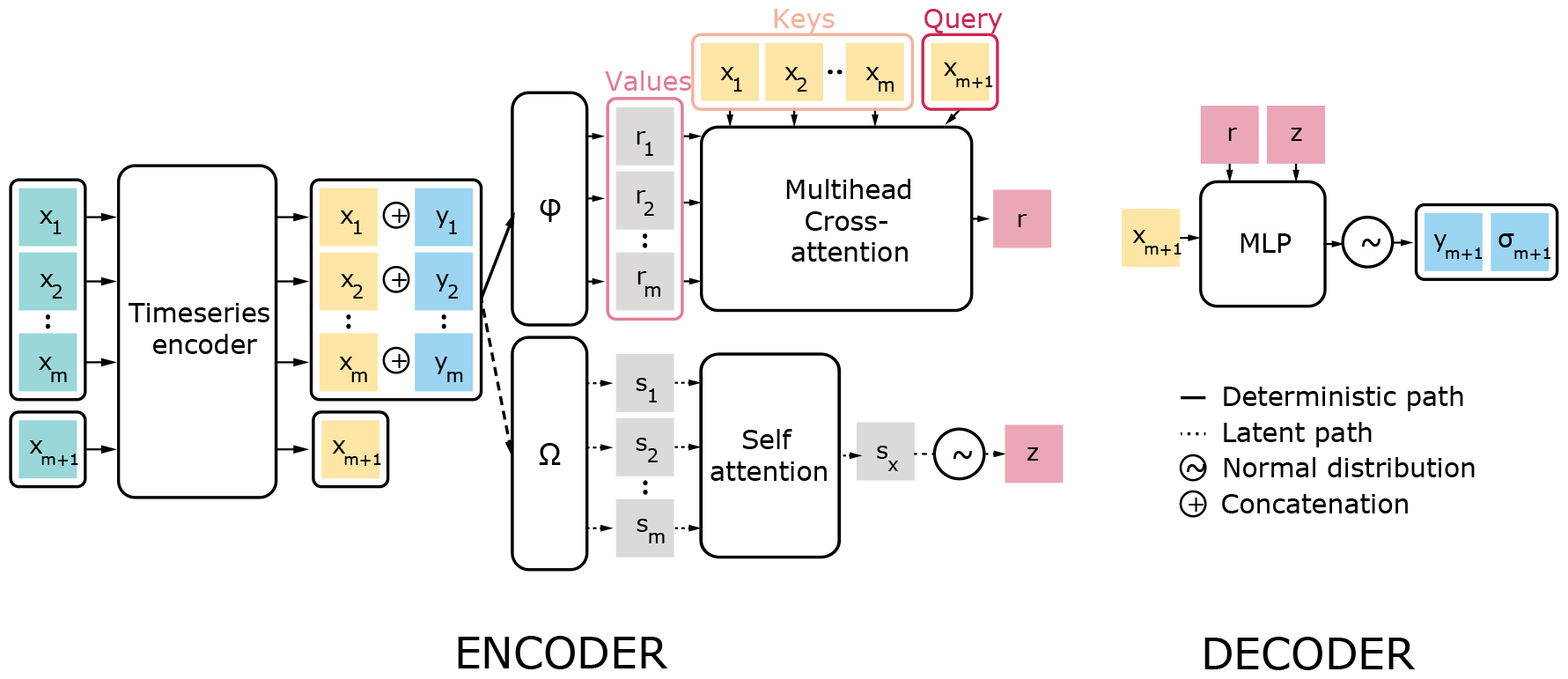
Attentive Neural Processes architecture for timeseries

### 2.7 Objective/Loss Function

#### 2.7.1 MSE Loss

The objective function for RNN and transformer model variations used mean squared error measuring the discrepancy between patient’s reference outcome values <u>**y**_*i*_</u> and its corresponding predicted outcome values <u>**ŷ**_*i*_</u> in the training dataset Eq. 1.

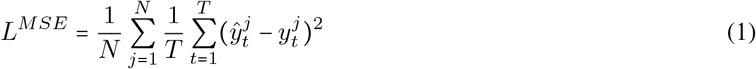

#### 2.7.2 ANP Loss

For the ANP model variations, the model parameters were optimized by maximizing the evidence lower bound (ELBO). This translates to maximizing the loglikelihood of the outcome targets and minimizing the Kullback–Leibler divergence (i.e. relative entropy) between the computed summaries of the targets and the contexts Eq. 2. For a training set with *N* samples, for each sample, a random set of contexts and targets are generated, a sample loss is computed and then averaged across all samples to compute total ANP loss Eq. 3. Lastly, we experimented with a mixed loss that is a convex combination between MSE and ANP loss using *γ* ∈ [0,−1].

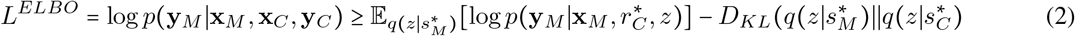

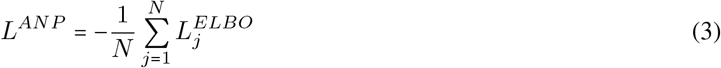

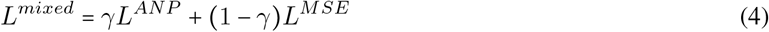

An *l*_2_-norm regularization term *λ* (i.e. hyperparameter) applied to the model weights and was used in all the loss formulations. The training was done using mini-batches, computing the loss function and updating the parameters/weights occurred after processing each mini-batch of the training set.

### 2.8 Baseline non-sequential models

For baseline models, we trained linear regression models such as Ridge, Lasso, and ElasticNet, and tree-based regression models such as Random Forest, Histogram-based Gradient Boosting Tree, and eXtreme Gradient Boosting (XGBoost).

## 3 Experimental setup

We followed a stratified 5-fold cross-validation scheme, in which the dataset was split into 5 folds, each having a training and test set size of 80% and 20% of the data, respectively, and a validation set size of 10% of the training set in each fold (used for hyperparameter selection in case of neural models). For each fold, a model was trained on the training sequences and then evaluated on the corresponding test sequences of that fold. Model performance was evaluated using root mean squared error (RMSE), and weighted RMSE (which corresponds to computing RMSE for each patient separately and then taking average across all patients). The weighted RMSE metric is analogous to *length-wise* weighting where the model performance is captured across wide range of sequence lengths (especially for shorter sequences). During models’ training, the epoch in which the model achieved the best harmonic mean between both scores on the validation set was recorded, and model state as it was trained up to that epoch was saved. This best model, as determined by the validation set, was then tested on the test split. The evaluation of the trained models was based on their average performance on the test sets across the 5 folds.

### 3.1 Hyperparameter search

We used a uniform random search strategy [34] that randomly chose a set of hyperparameters configurations (i.e. embedding dimension, number of hidden layers, dropout probability, etc.) from the set of all possible configurations and trained corresponding models on each fold using their respective training and validation data only. Then the best configuration for each model (i.e. the one achieving best performance on the validation set) was used for the final training and testing for the corresponding fold. The range of possible hyperparameters configurations (i.e. choice of values for hyperparameters) for trained models is reported in Appendix D.

For baseline models, we used random search strategy over each model’s specific hyperparameter space using 2-fold cross validation on the combined training and validation set of each fold of the 5 folds separately. Then the model achieving best performance is retrained and tested on each of the the corresponding fold.

### 3.2 Uncertainty quantification

We denote the lower and upper uncertainty prediction for an *i*−th patient at time *t* by 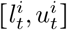 respectively. A model might estimate outcome uncertainty in terms of the variance (or standard deviation 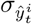) of its generated prediction 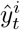 to create an interval 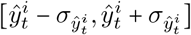. Hence, we can evaluate the generated interval using (a) coverage, defined by the average number of times the true outcome 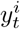 lies within the predicted uncertainty interval, and (b) winkler score [35], defined by Eq. 5 that evaluates the average width of the prediction interval (i.e. the *tightness* of the uncertainty estimate or prediction interval) while penalizing incorrect predictions outside the interval. The score is computed for all prediction events for the patients in the test set to get an overall score.

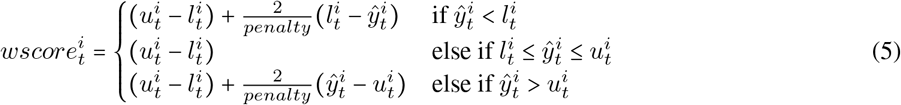

For ANP RNN models, we used the estimated uncertainty (i.e 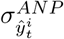 Eq. 38) to construct the prediction intervals and compute the coverage and winkler scores. For RNN models, we used Monte Carlo Dropout (MCDropout) [36] where during inference time, a model ran for multiple rounds with dropout layers activated (i.e. dropping out randomly neurons) and a prediction is made for each round. Then we computed the average 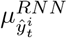 and standard deviation 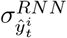 of these predictions to build a predictive distribution. The process of dropping out neurons is analogous to creating a new variant of the model/architecture where the prediction of this network is considered as a new sample from the space of models corresponding to different dropout configurations.

Additionally, we used split conformal prediction [23] to improve the uncertainty estimates computed by the RNN and ANP RNN models by turning them into rigorous prediction intervals with certain coverage guarantees. The idea is to compute non-conformity scores using the validation data quantifying the error (i.e. distance) between the prediction and the true outcome 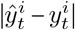, and weighted by the inverse of the uncertainty 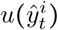 estimated by the models (i.e. 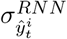 and 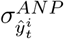). Then we determine 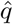 to be the empirical 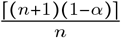 quantile of the non-conformity scores (i.e. computed using the validation data) where 1 − *α* is the desired coverage. We chose *α* = 0.1 (i.e. aimed for coverage≥ 90%) and the updated prediction intervals will be 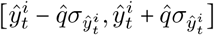 where the prediction 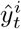 and standard deviation 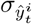 are computed by the evaluated models (i.e RNN and ANP RNN). We then computed the coverage and winkler scores evaluating the newly updated prediction intervals.

### 3.3 Post-hoc explainability

#### 3.3.1 Patient similarity based explainability

We evaluated the usefulness of the computed latent representations from our ANP model (i.e. computed vector representation) to retrieve similar patients. Given a patient journey/history representation at a prediction time-point, we computed distances (such as *L*^1^, *L*^2^, and *cosine*) to all other representations and selected the *k* closest patient embeddings.

We matched the computed patient representations from the test set to their closest representations in the train set, such that we found the subset of nearest neighbour representations. Then using *k*−NN regression, we compared the representation’s future FVC% with the average FVC% of their closest matched set. We evaluated the prediction performance in two cases when using *k*−NN with (a) the computed representation from the ANP model and (b) the raw input features.

#### 3.3.2 Feature importance derived from similarity assessment

Once we established the “predictive utility” of summarizing patient’s timeline using the computed latent representation from the ANP model, we inspected the role of each raw input feature in the similarity computation between an index patient and their subset of nearest neighbours’ latent representations.

For continuous features, we computed the average absolute distance (AAD) between the input feature values of the patients in the test set (ℛ_*test*_) and the average value in their matched set 𝒩 _*e*_ (in the training data). In this setup, the raw input features represent a running average of the input features across the past visits in the trajectory up to the event/visit where we predict the future FVC outcome. We also computed a modified version denoted by the standardized AAD, dividing the AAD by the standard deviation of the feature:

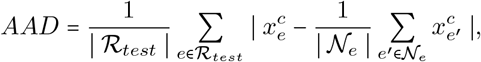

Where 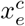 is the value of the continuous feature *c* for patient embedding vector *e*. This average distance quantifies the average deviation of the feature values in the nearest neighbours subset from the feature values of the index patient. The smaller the distance is, the bigger the influence/contribution of the feature in the similarity computation.

Similarly for categorical features, we computed the average absolute distance between the categorical input feature values of the reference patients in the test set ((ℛ_*test*_) and the average value in their matched set 𝒩_*e*_ (in the training data). A major difference with respect to the continuous case is that we only considered available (i.e. present) features for each reference patient when performing the computation. This allows to distinguish between the features that are commonly present and influential in the similarity computation (i.e. has small distance) from the ones that are frequently not present.

## 4 Results

Patients’ FVC% measures ranged from 22 to 150% predicted with a mean (SD) of 90.53% predicted (21.52). Overall, sequential neural models showed better performance compared to tree- and regression-based models where RNN and ANP RNN (both versions) achieved best performance with average (SD) 8.243 (0.185), 8.240 (0.168) weighted RMSE, and 6.935 (0.211), 6.94 (0.190) MAE, respectively (Table 4). Lasso and Histogram-based gradient boosting regressor were best among baseline models with average 8.479 (0.201), 8.524 (0.329) weighted RMSE, and 7.121 (0.219), 7.173 (0.386) MAE respectively. In comparison, a naïve baseline using the mean FVC% predicted value as a predictor would achieve 18.718 (0.317) weighted RMSE, and 17.619 (0.599) MAE. Then we used the ANP RNN model predictions along the learned latent embedding as input to two separate logistic regression models trained to predict an FVC% decline and FVC% increase of at least 10% points and achieved an average of 0.704 and 0.70 AUC scores respectively across the 5-folds.

**Table 4:**
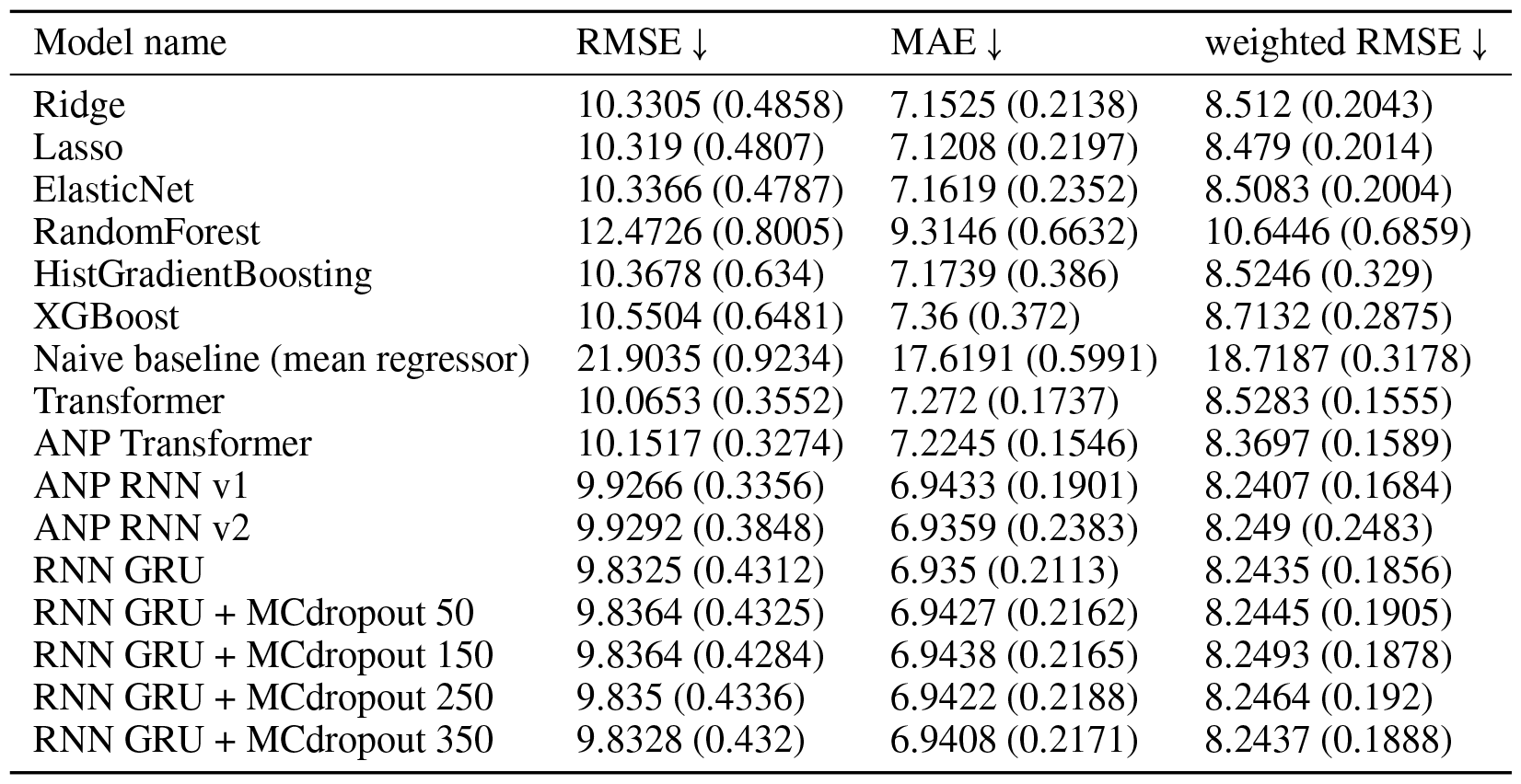
Models’ average performance across 5-folds.

| Model name | RMSE ↓ | MAE ↓ | weighted RMSE ↓ |
| --- | --- | --- | --- |
| Ridge | 10.3305 (0.4858) | 7.1525 (0.2138) | 8.512 (0.2043) |
| Lasso | 10.319 (0.4807) | 7.1208 (0.2197) | 8.479 (0.2014) |
| ElasticNet | 10.3366 (0.4787) | 7.1619 (0.2352) | 8.5083 (0.2004) |
| RandomForest | 12.4726 (0.8005) | 9.3146 (0.6632) | 10.6446 (0.6859) |
| HistGradientBoosting | 10.3678 (0.634) | 7.1739 (0.386) | 8.5246 (0.329) |
| XGBoost | 10.5504 (0.6481) | 7.36 (0.372) | 8.7132 (0.2875) |
| Naive baseline (mean regressor) | 21.9035 (0.9234) | 17.6191 (0.5991) | 18.7187 (0.3178) |
| Transformer | 10.0653 (0.3552) | 7.272 (0.1737) | 8.5283 (0.1555) |
| ANP Transformer | 10.1517 (0.3274) | 7.2245 (0.1546) | 8.3697 (0.1589) |
| ANP RNN v1 | 9.9266 (0.3356) | 6.9433 (0.1901) | 8.2407 (0.1684) |
| ANP RNN v2 | 9.9292 (0.3848) | 6.9359 (0.2383) | 8.249 (0.2483) |
| RNN GRU | 9.8325 (0.4312) | 6.935 (0.2113) | 8.2435 (0.1856) |
| RNN GRU + MCdropout 50 | 9.8364 (0.4325) | 6.9427 (0.2162) | 8.2445 (0.1905) |
| RNN GRU + MCdropout 150 | 9.8364 (0.4284) | 6.9438 (0.2165) | 8.2493 (0.1878) |
| RNN GRU + MCdropout 250 | 9.835 (0.4336) | 6.9422 (0.2188) | 8.2464 (0.192) |
| RNN GRU + MCdropout 350 | 9.8328 (0.432) | 6.9408 (0.2171) | 8.2437 (0.1888) |

When comparing uncertainty estimates from the models predictions (Table 5), ANP RNN models achieve up to 79% coverage on average compared to RNN models with Monte Carlo dropout (for varying number of runs) with an average of 17.5% (Figure 8a). A large difference between both models is also observed when comparing their winkler scores (smaller score is better) that take into consideration the average length of the prediction interval (i.e. uncertainty estimates) while penalizing incorrect predictions outside the interval. Adding conformal prediction (i.e. conformalizing the models’ prediction and uncertainty estimate scores), with *α* = 0.1 (i.e. aimed for coverage ≥ 90%), all models achieved the desired coverage while ANP RNN still having the lowest winkler score (Table 5 and Figure 8b). Though, the gap between RNN and ANP RNN winkler scores is drastically reduced when using conformal prediction procedure.

**Table 5:**
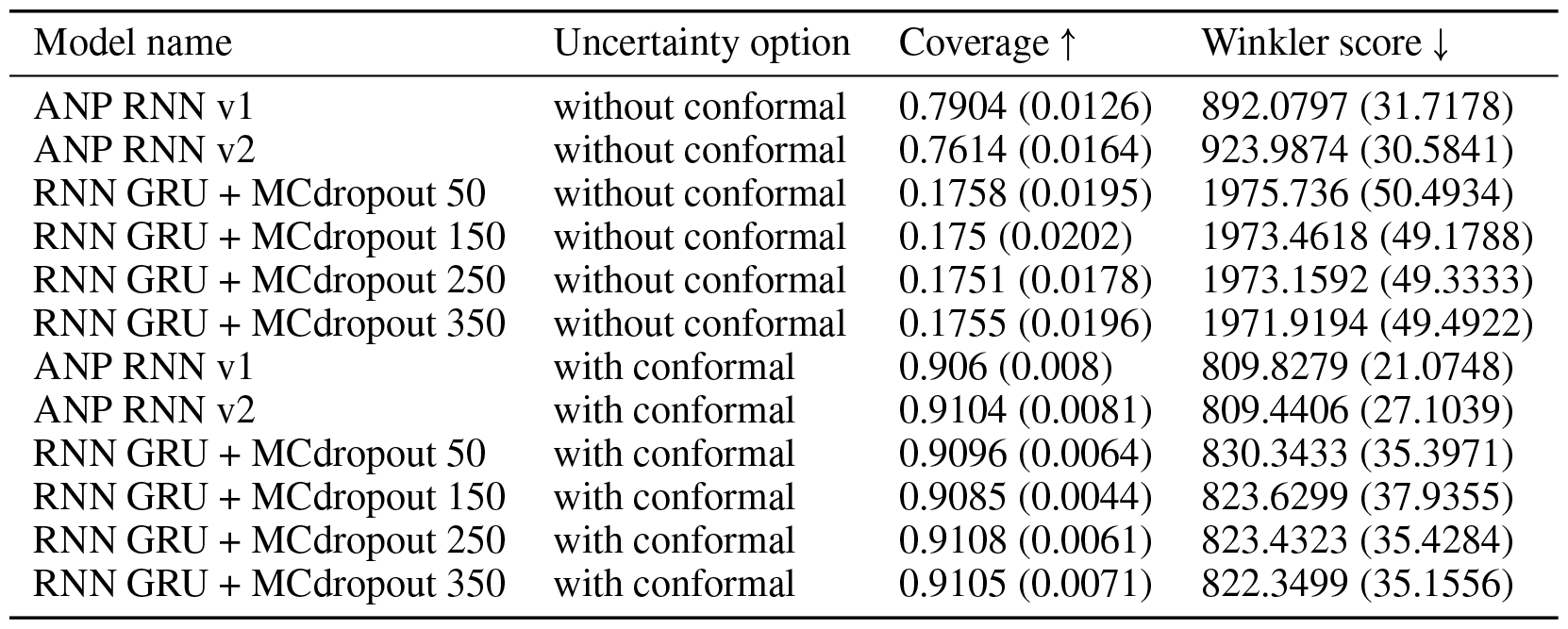
Evaluation of models’ uncertainty prediction across 5-folds.

| Model name | Uncertainty option | Coverage ↑ | Winkler score ↓ |
| --- | --- | --- | --- |
| ANP RNN v1 | without conformal | 0.7904 (0.0126) | 892.0797 (31.7178) |
| ANP RNN v2 | without conformal | 0.7614 (0.0164) | 923.9874 (30.5841) |
| RNN GRU + MCdropout 50 | without conformal | 0.1758 (0.0195) | 1975.736 (50.4934) |
| RNN GRU + MCdropout 150 | without conformal | 0.175 (0.0202) | 1973.4618 (49.1788) |
| RNN GRU + MCdropout 250 | without conformal | 0.1751 (0.0178) | 1973.1592 (49.3333) |
| RNN GRU + MCdropout 350 | without conformal | 0.1755 (0.0196) | 1971.9194 (49.4922) |
| ANP RNN v1 | with conformal | 0.906 (0.008) | 809.8279 (21.0748) |
| ANP RNN v2 | with conformal | 0.9104 (0.0081) | 809.4406 (27.1039) |
| RNN GRU + MCdropout 50 | with conformal | 0.9096 (0.0064) | 830.3433 (35.3971) |
| RNN GRU + MCdropout 150 | with conformal | 0.9085 (0.0044) | 823.6299 (37.9355) |
| RNN GRU + MCdropout 250 | with conformal | 0.9108 (0.0061) | 823.4323 (35.4284) |
| RNN GRU + MCdropout 350 | with conformal | 0.9105 (0.0071) | 822.3499 (35.1556) |

To evaluate the utility of the summary (i.e. latent representation) computed by our ANP, we used *k*−NN regression on the latent embeddings model for the patient’s journey/trajectory by comparing the future FVC% values of the embeddings in the test set with the average values of their most similar embeddings, as computed by *k*−NN regression on the latent embeddings. We further compared the performance of our approach to the performance of a *k* − NN algorithm applied to the raw data. The *k*-NN model on the latent representations was a clear winner achieving better performance compared to using raw input features (see Figure 4a vs. Figure 4b).

**Figure 4:**
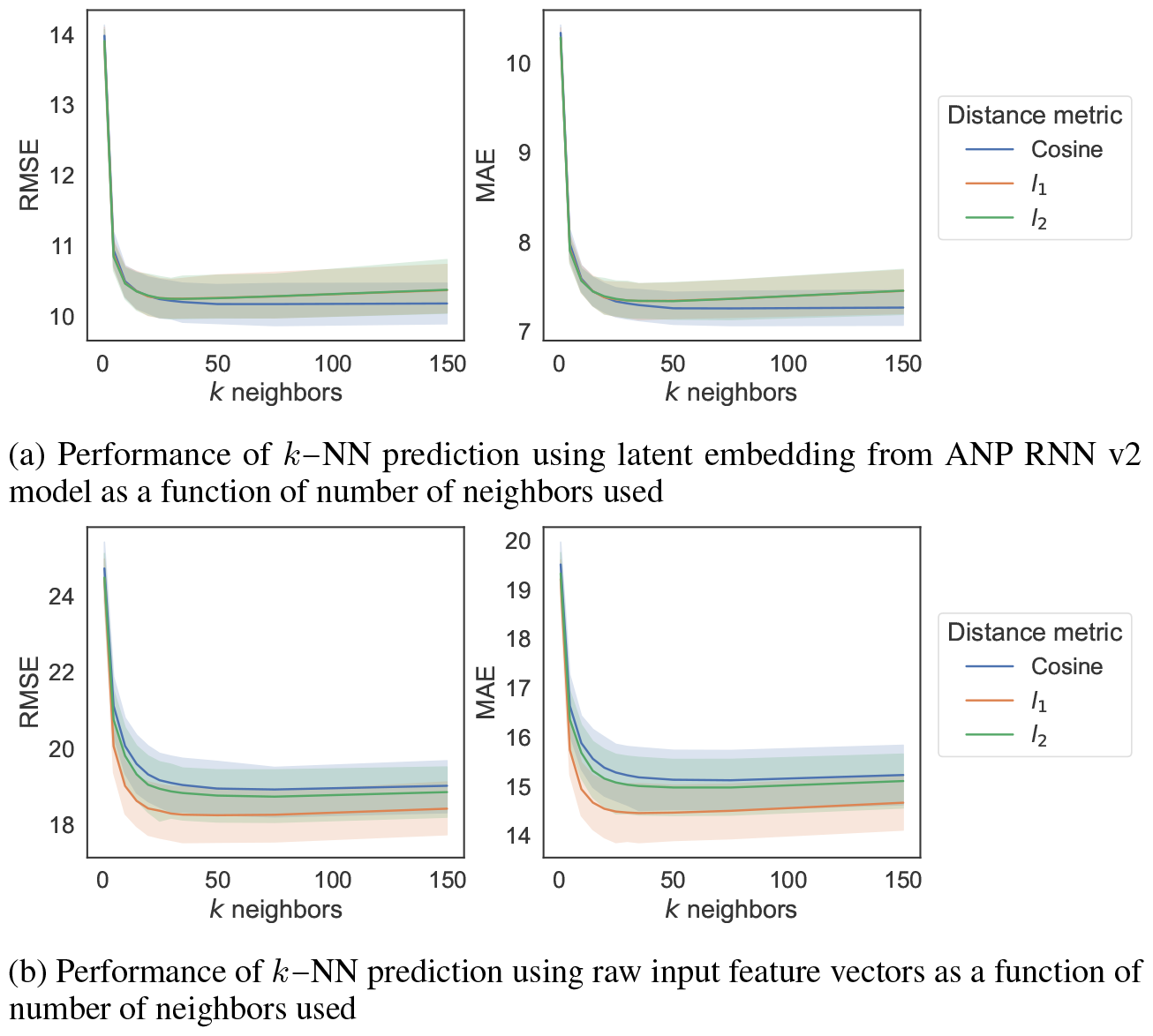
*k*−NN regression using latent embedding vs. raw input features

We then investigated the models’ features attribution (i.e. importance) using the SHAP scores from the trained LASSO models (i.e. best baseline models) across the five folds. The features were ranked based on their importance from the top-10 features across the five trained models (i.e. based on 5-folds), then averaged and reported in Figure 5a. Multiple related features (such as categorical ones) were joined using the sum of their SHAP scores. Previous measurement of FVC% predicted, extent of skin involvement, previous measurement of DLCO%, therapies (documented and missing indicators), anti-centromere positivity, age, dyspnea, CRP elevation and digital ulcers were the top-10 consistent features across the 5-folds. Inspecting the main contributing features, previous FVC and DLCO values (i.e. relative to the next prediction event), were positively correlated with the SHAP scores indicating higher previous values had positive influence and lower previous values had negative influence on future prediction of FVC values respectively Figures 5b, 5c. Patients being diagnosed with diffuse and limited cutaneous skin involvement had a negative influence (based on their SHAP scores) on the prediction of future FVC values in contrary to patients with no skin involvement Figure 5d. Prescribed therapies such as methotrexate, chloroquine/hydroxychloroquine, mycophenolic acid, and rituximab have a positive influence on the prediction of next FVC value Figure 5e. Furthermore, being diagnosed with significant dyspnea or dyspnea NYHA 3 have negative influence on the prediction of future FVC values in contrary to not having significant dyspnea or being diagnosed with a first stage NYHA dyspnea Figures 5g, 5i. Lastly, having no CRP elevation affects positively the prediction Figure 5h in contrary to having digital ulcers that affects negatively the prediction of future FVC scores Figure 5f.

**Figure 5:**
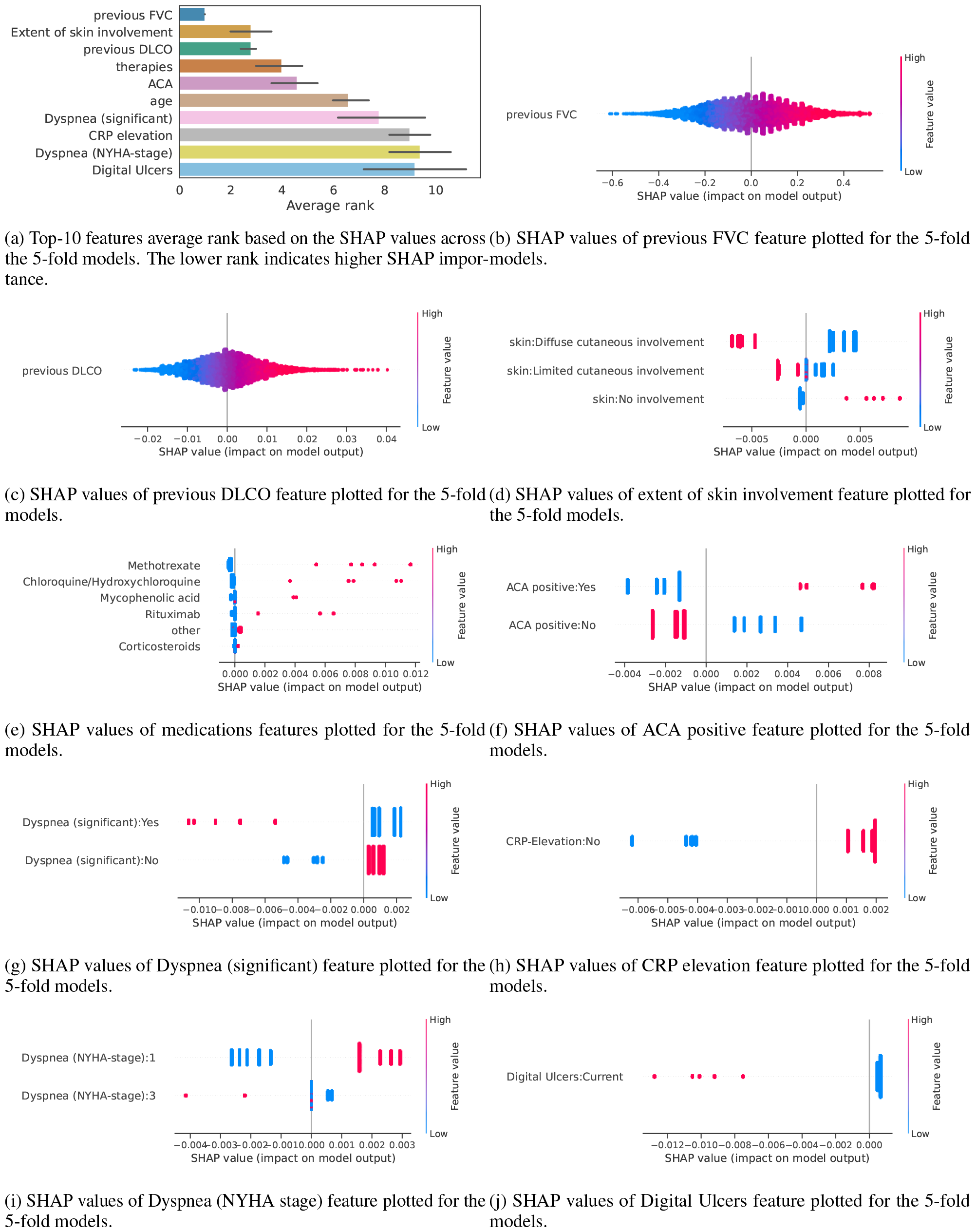
Feature importance using SHAP scores for the 5-fold models

Following our similarity-based analysis to compute feature importance as described in subsubsection 3.3.2, Figure 9 provides insights into the nearest neighbour attribution mechanism at the patient level. The features are ranked from the most to least important for both the continuous and categorical features. Overall, the top features overlap with the SHAP analysis highlighting the importance of previous FVC% and DLCO% values along with time difference between events and time to prediction. Moreover, using the patient similarity approach, we can inspect and visualize the characteristics (i.e. features) of each reference patient and their nearest neighbors from the training set as shown in both Figures 12 and 13. We can identify the top most-similar features between the reference patient and nearest neighbors in addition to providing the FVC% predictions from the neural model, k-NN model and all FVC% values from nearest neighbors.

We further inspected the latent vector representation *z* computed by the ANP RNN model for every prediction event in patients’ trajectories (from the test sets), and visualize it using tSNE [37] (embedding the vectors in ℝ ^2^) to become points with 2D coordinates Figure 6. Then we highlighted each point (i.e. representing the tSNE embedding of *z*) with different annotations corresponding to the main features describing the event such as current DLCO value, dyspnea status, ACA status, extent of skin involvement, time elapsed from the first event of a patient’s trajectory, and the predicted future FVC% predicted value. Overall, we see an agreement with the SHAP attribution analysis of the baseline model, where patients predicted with higher FVC values had higher current DLCO values and/or ACA positive status, and patients predicted with lower FVC values had significant dyspnea and/or diffuse cutaneous skin involvement and the inverse was true.

**Figure 6:**
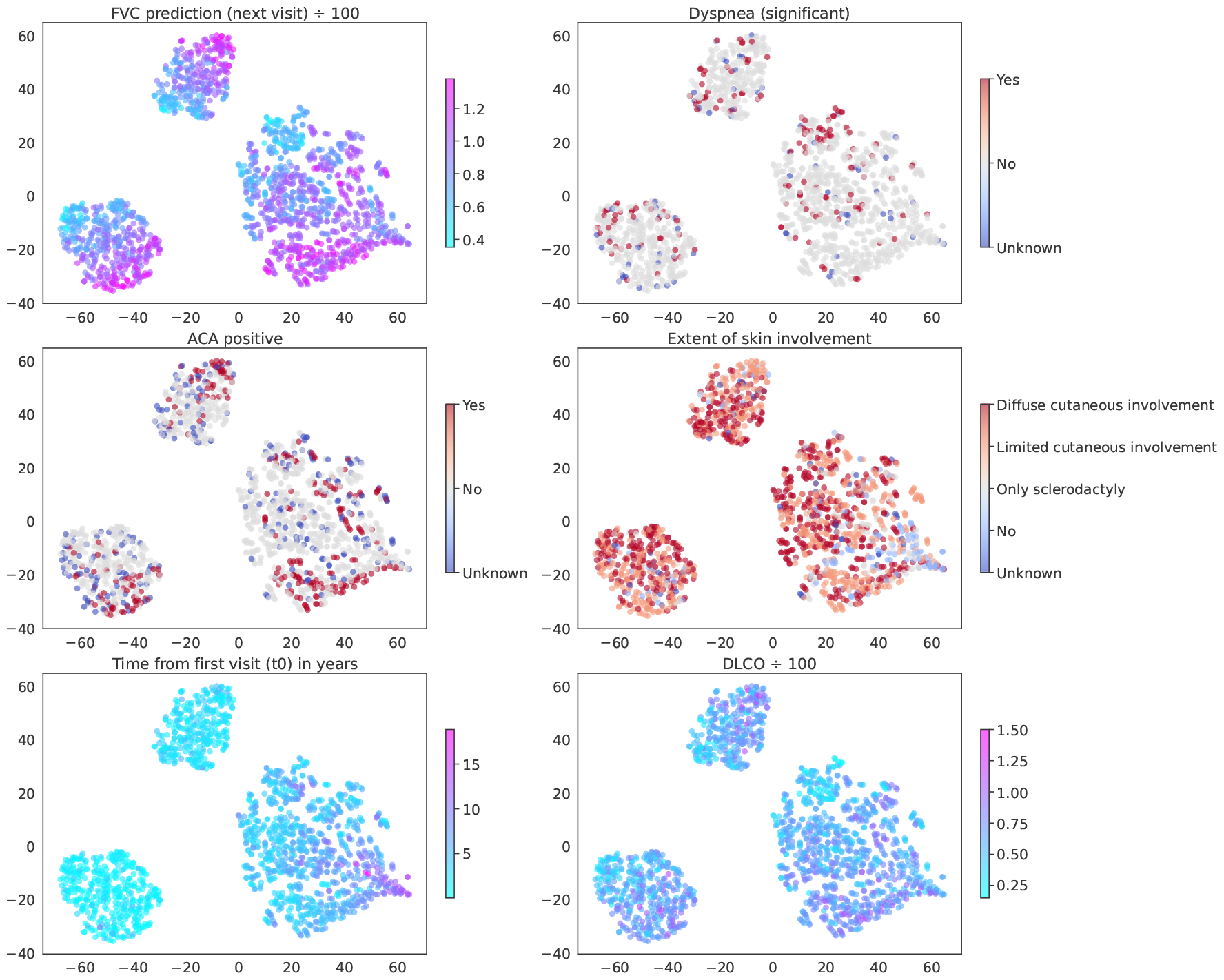
tSNE embedding of latent path vector *z* from ANP RNN v2 model computed for every future prediction event in one of the test set folds.

Lastly, we analyzed the best sequential models’ performance (i.e. RNN and ANP RNN) as a function of therapy documentation (i.e. defined as the percentage of visits in a patient’s trajectory that had documentation about therapy - i.e. not missing) and coverage (percentage of visits that had present (available) therapies in a patient’s trajectory). A therapy documentation or coverage ≥ 0 means we computed the models’ performance using all patients, and as we incrementally increased the therapy documentation or coverage criterion, we evaluated the models’ performance on patients with higher ratio of therapy. We observed on average a decreasing trend in RMSE and weighted RMSE indicating better performance with more comprehensive therapy documentation history (Figures 7 and 10).

**Figure 7:**
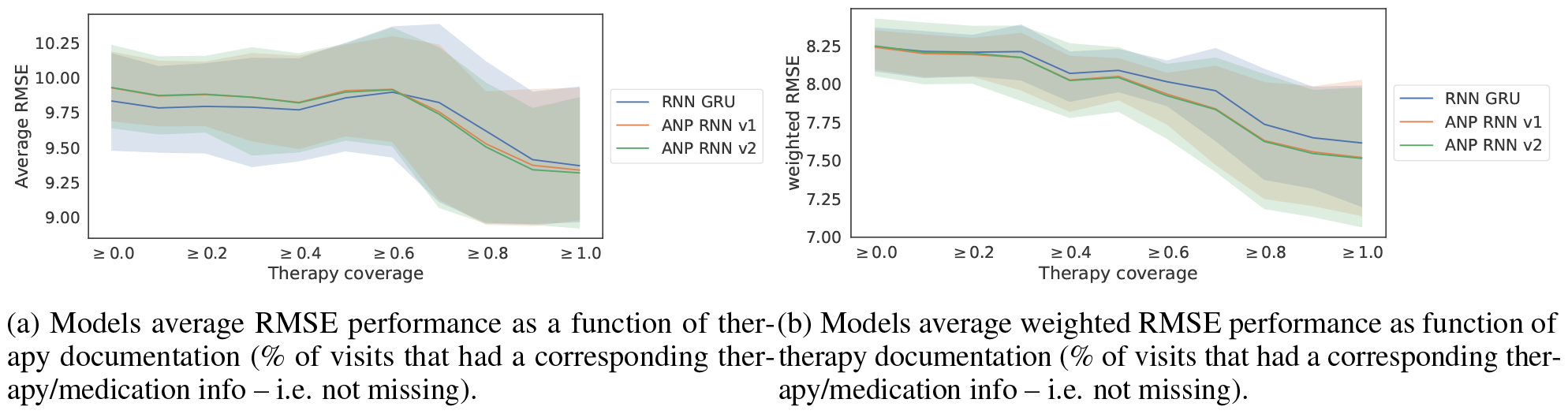
Performance versus therapy documentation

**Figure 8:**
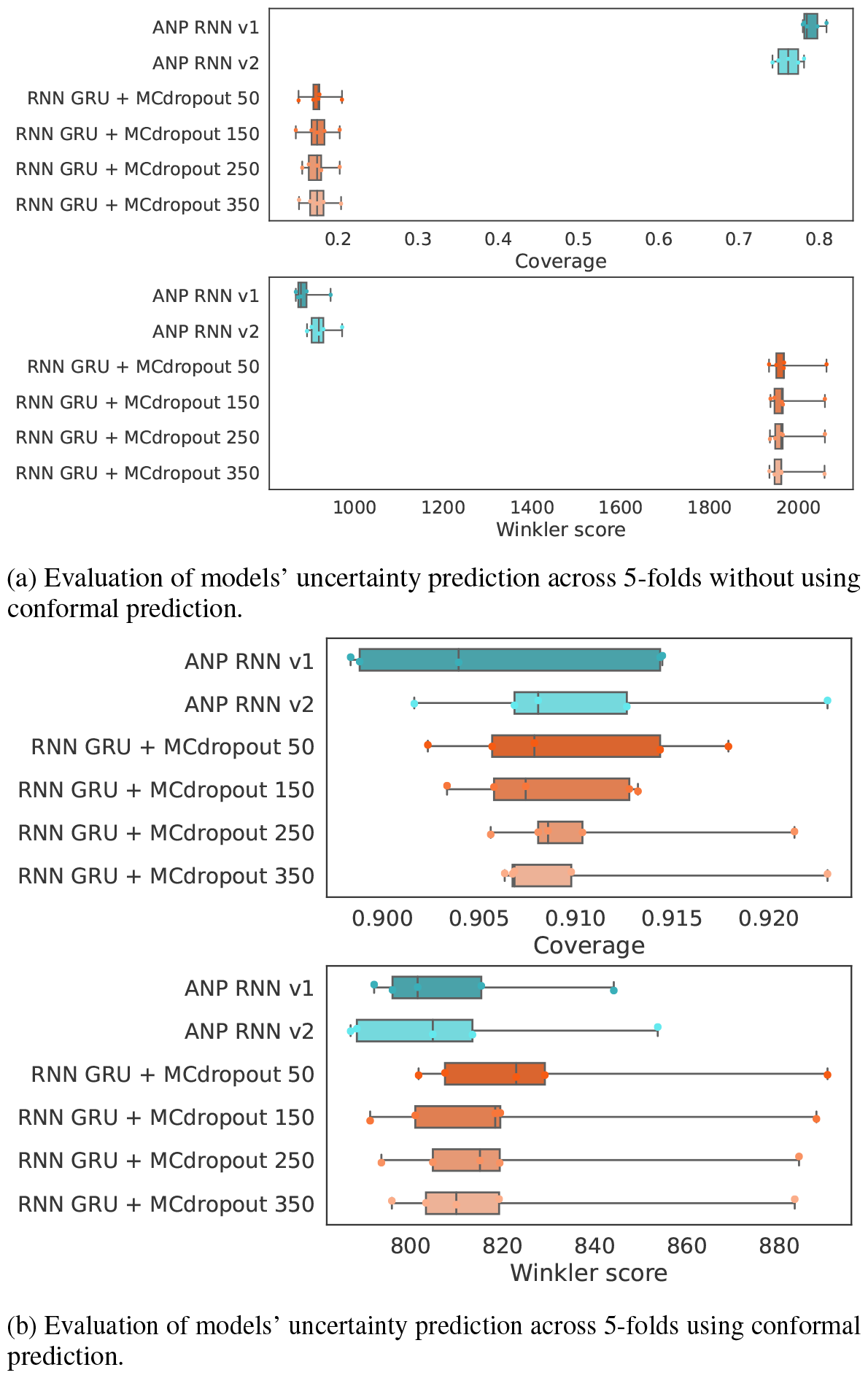
Models uncertainty evaluation with and without conformal prediction.

**Figure 9:**
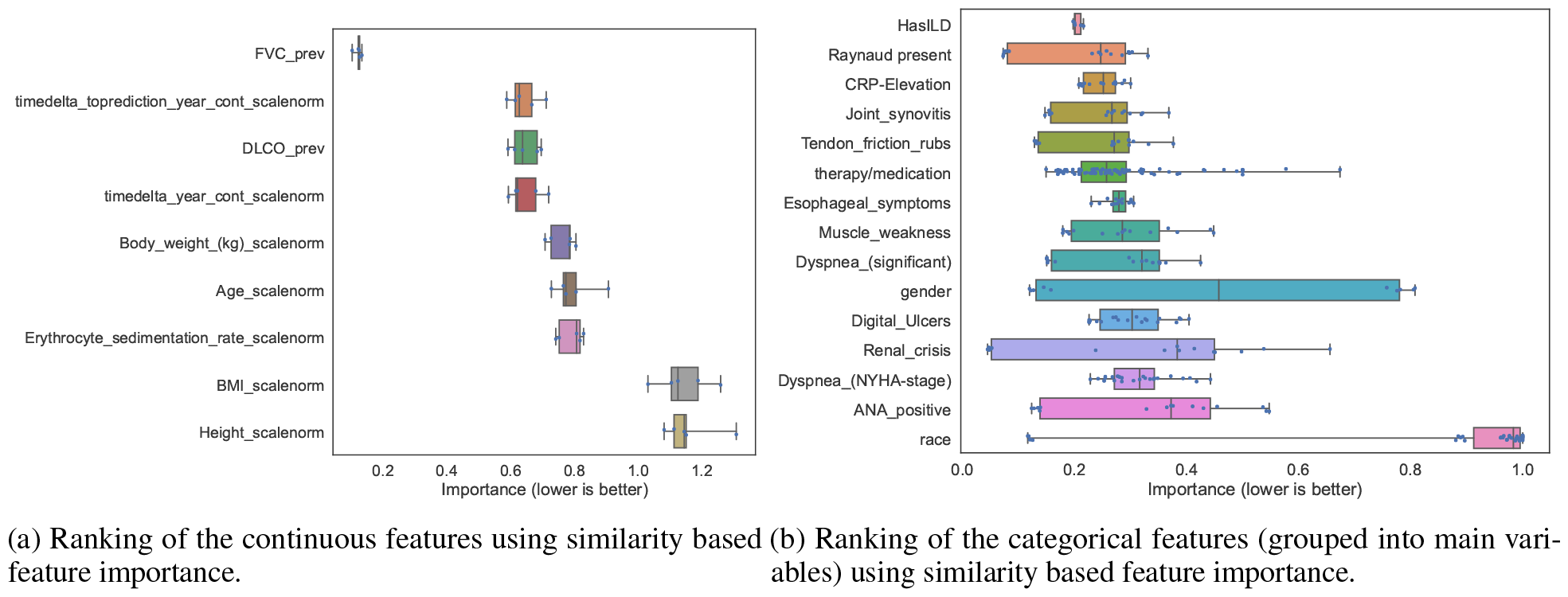
Feature importance computation using similarity assessment.

**Figure 10:**
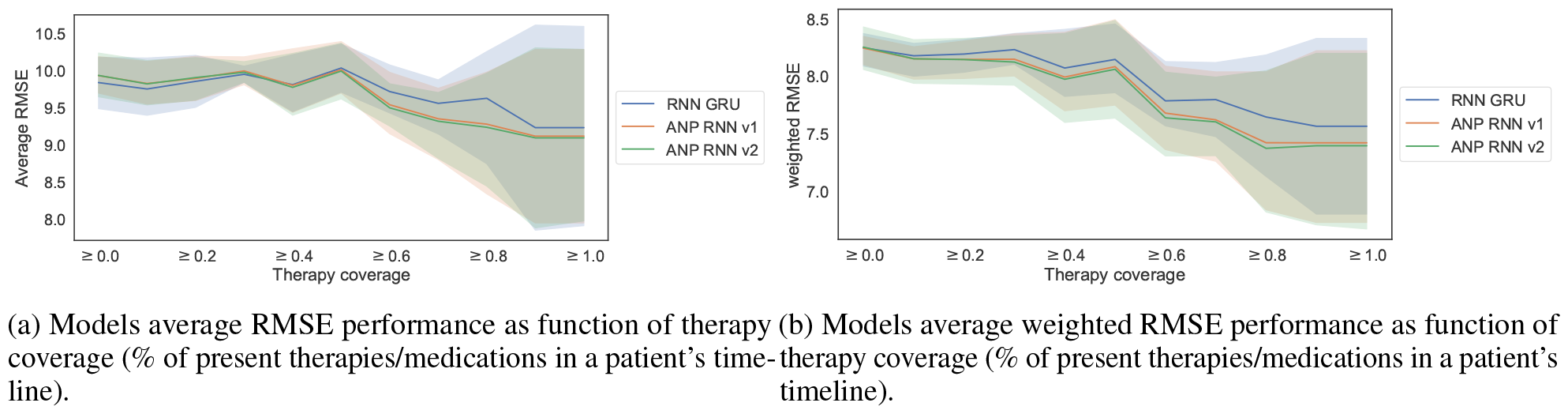
Performance versus therapy coverage

**Figure 11:**
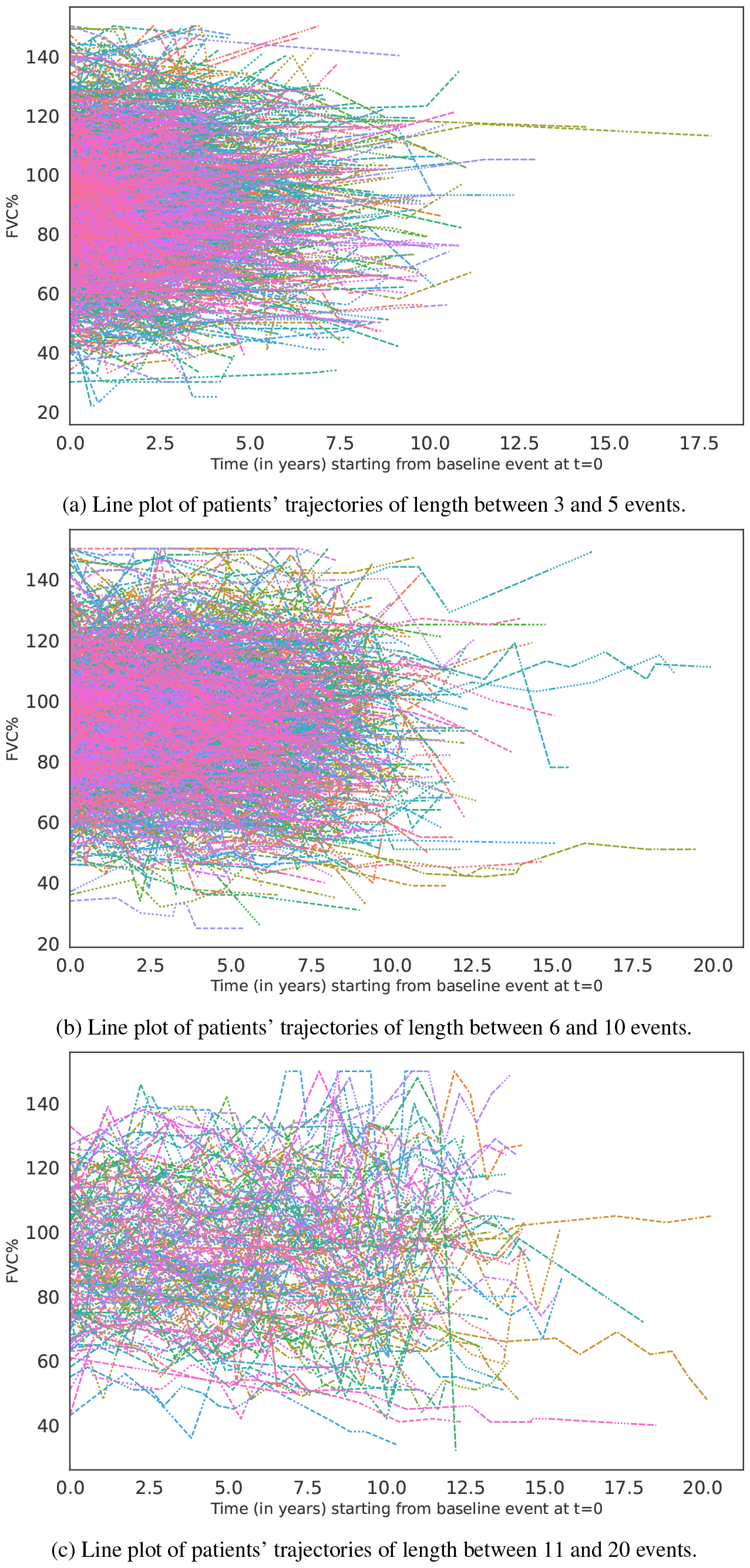
Line plot of patients’ trajectories stratified by number of events in timeline.

**Figure 12:**
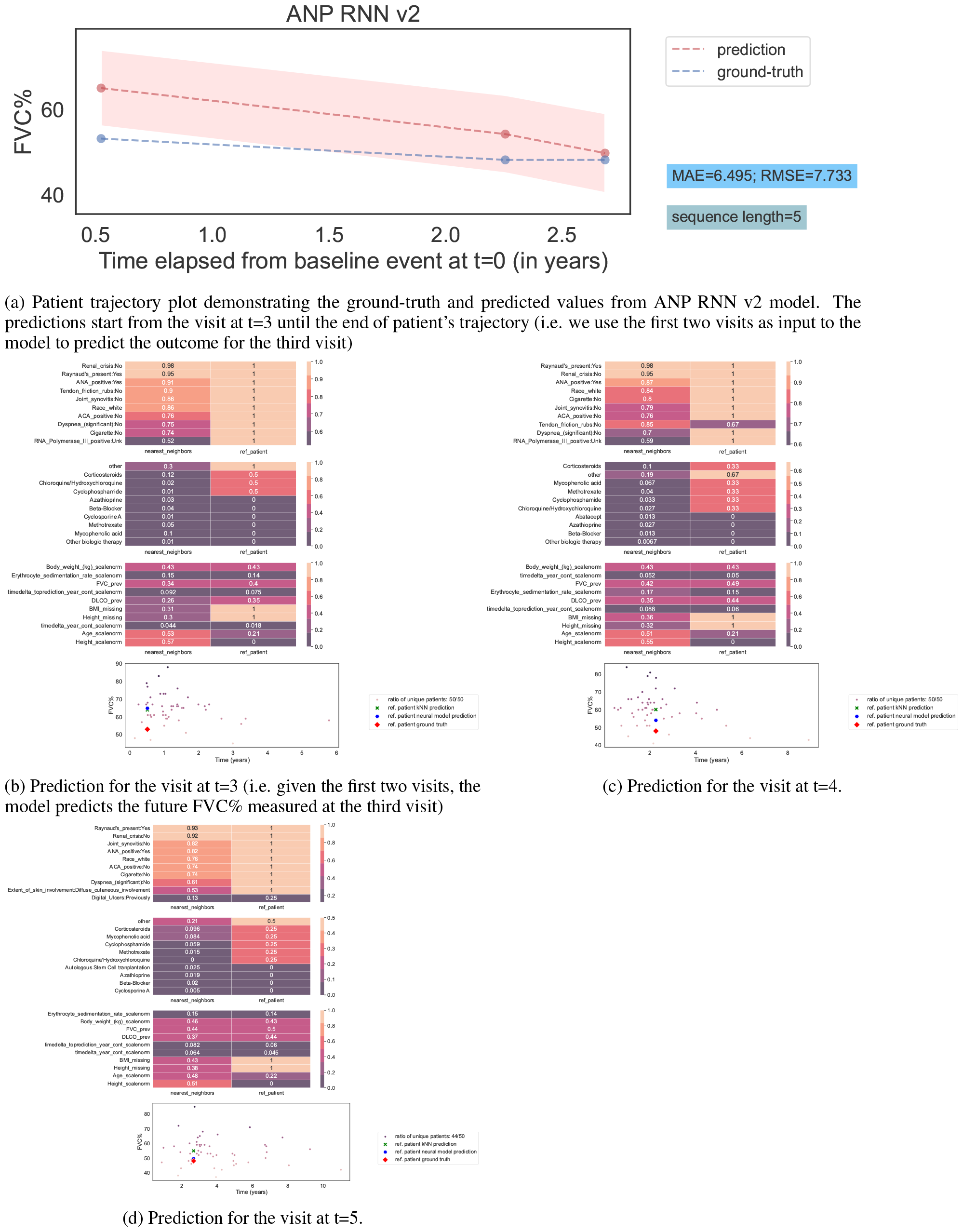
Patients’ timeline and prediction at each visit/event.

**Figure 13:**
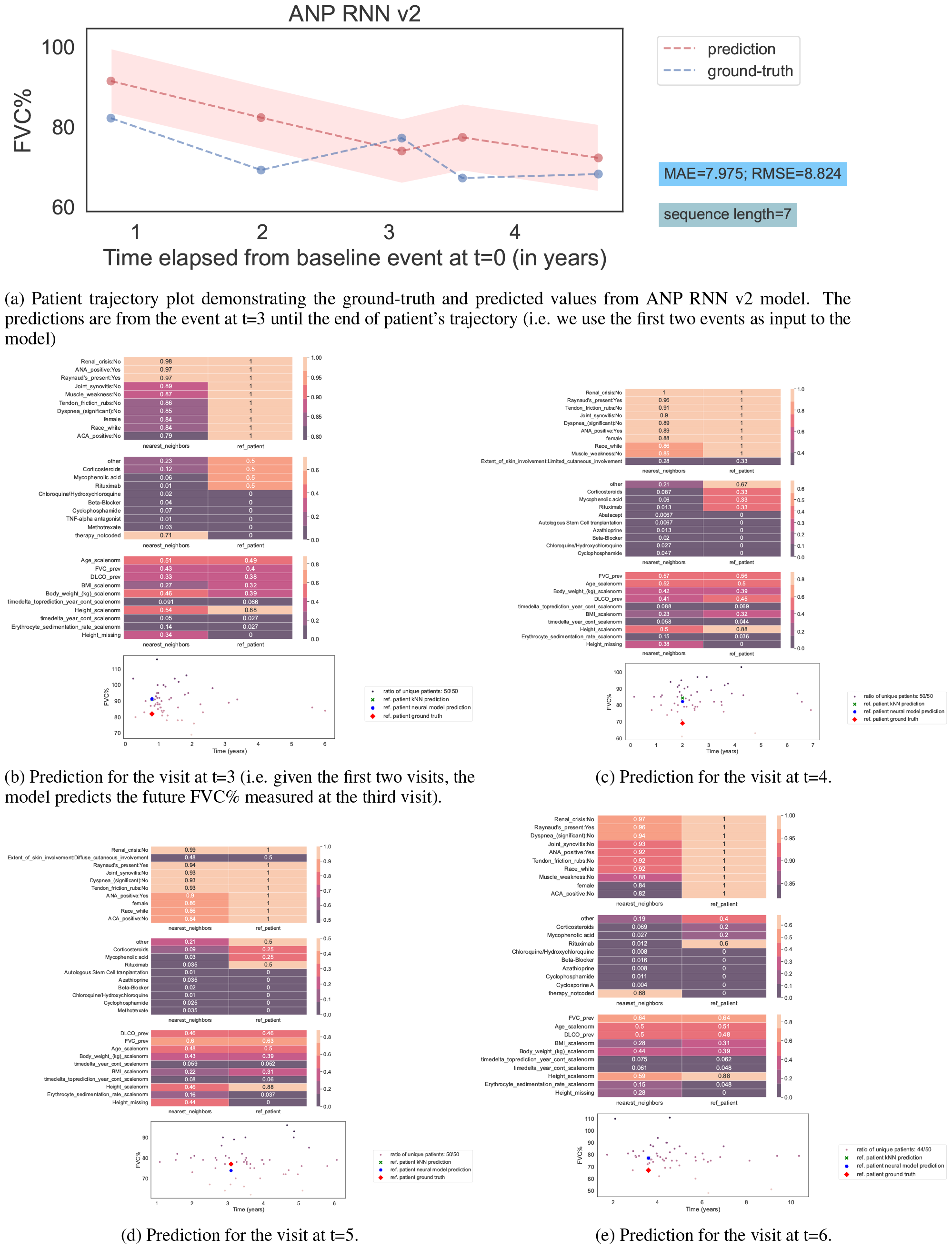
Patients’ timeline and prediction at each event.

## Discussion

Our study demonstrates the feasibility of predicting SSc-ILD patients’ future FVC values, i.e. lung function trajectories, on an individual base using machine learning. Overall, sequential models achieved the best performance compared to tree-based and regression models, with recurrent neural network and attentive neural process variations having the lowest RMSE, MAE and weighted RMSE scores. These models would need only one baseline measurement (in case of RNN) and two measurements (in case of ANP) as input to start predicting the future FVC% values. Moreover, our study suggests that attentive neural process based models can provide uncertainty quantification on the predicted outcomes out of the box with an average coverage of 79%. In contrast, running RNN based models with Monte Carlo dropout achieved little less than 18% coverage. As it is a common approach to use Monte Carlo dropout [36] with trained neural network models to provide a notion of uncertainty, we show that this is suboptimal compared to using uncertainty estimates from a trained attentive neural process model. Furthermore, using split conformal prediction [23] to adjust the uncertainty estimates from both models proved to be a viable strategy, achieving the desired coverage while generating tighter uncertainty estimates. This provides additional evidence for the efficacy of using conformal prediction as a post-hoc model-independent procedure for creating uncertainty estimates on the predicted outcomes.

The analysis of feature importance (or feature attribution) in the model’s prediction revealed the main features that align with the literature on the potential risk factors for predicting ILD progression. Similar to previous findings, our analysis also indicates that for the prediction of FVC% predicted values, the main features include previous FVC and DLCO measures [38, 39, 13], as well as other factors such as extent of skin involvement, age, CRP elevation, and dyspnea being among the top 10 identified predictors [12, 13, 38, 40, 41]. Moreover, our analysis identified additional features such as the presence of ACA, digital ulcers and immunomodulating therapies that play a role in the prediction of future FVC values. These features were also highlighted when visualizing the latent representation computed by the attentive neural process model showing that compressing the patient trajectory into this representation is useful for the prediction of future FVC values. In addition, computing patient similarity allowed us to provide additional inspection mechanisms to demonstrate the overall patient trajectory and the visit level characteristics of the reference patient and their corresponding nearest neighbors. This information helps to offer additional insight between input features and the predicted outcome while giving the physicians access to multiple outcome possibilities (corresponding to nearest neighbor future FVC% predicted values) in addition to the trained model’s prediction.

Lastly, we showed the importance of therapy documentation/coverage and its effect on improving models’ prediction performance. Having patients with more detailed therapy documentation, a model can further use this information to improve its prediction. Thus, it is important to have accurate and complete documentation of therapies in healthcare datasets if we aim to build better prediction models in the future.

## Data Availability

The raw dataset is owned by the EUSTAR group, and may be obtained by request after the approval and permission from EUSTAR board.

https://eustar.org/

## Availability of data and materials

The raw dataset is owned by the EUSTAR group, and may be obtained by request after the approval and permission from EUSTAR board. The pre-processing scripts and the models’ implementation workflow will be made publicly available at https://github.com/uzh-dqbm-cmi/screener.

## Competing interests

The authors declare that they have no competing interests.

### Acknowledgements

The authors thank the patients and caregivers who made the study possible, as well as all involved clinicians from the EUSTAR who collected the data. This work is funded by the Swiss National Science Foundation (project number 201184).

## Collaborators

### EUSTAR collaborators (numerical order of centers)

2 Ulrich Walker, Universtitätsspital Basel, Dept. of Rheumatology, Basel, Switzerland; 4 Florenzo Iannone, Rheumatology DiMePReJ, School of Medicine, University Of Bari, Italy; 6 Oliver Distler, Department Of Rheumatology, University Hospital Zurich, Center For Experimental Rheumatology, University of Zurich, Zurich Switzerland; 7 Radim Be č vá ř, Institute of Rheumatology and Department of Rheumatology 1st Medical School, Charles University Na Slupi 4 (Prague) Czech Republic; 11 Maurizio Cutolo, Laboratory Of Experimental Rheumatology And Division Of Rheumatology Dept. Internal Medicine University Of Genova School Of Medicine, rccs San Martino Hospital, Genova, Italy; 13 Vasiliki Liakouli, Università della Campania, Naples, Italy; 16 Simona Rednic, Clinica Reumatologie, University Of Medicine And Pharmacy Iuliu Hatieganu Cluj, Emergency County Hospital Cluj, Cluj-Napoca, Romania; 17 Yannick Allanore, Rheumatology A Dpt, Paris 5 University Cochin Hospital, Paris, France; 23 Patricia E. Carreira, Rheumatology Department, Hospital Universitario 12 De Octubre, Madrid Spain; 25 Department Of Rheumatology And Immunology, Medical Centre, University Of Pécs, Pécs, Hungary; 28 Michele Iudici, Division Of Rheumatology, Geneva University, Hospitals Hôpital Beau-Séjour, Geneva, Switzerland; 31 Elisabetta Zanatta, Rheumatology Unit, Padua University Hospital, Padova, Italy; 35 Dominique Farge Bance, Department of Internal Medicine Hopital Saint-Louis, Paris, France; 38 Maria-Grazia Lazzaroni, Rheumatology And Clinical Immunology Unit, Asst Spedali Civili Of Brescia, University Of Brescia, Brescia, Italy; 40 Dirk Wuttge, Skane University Hospital – Lund, Lund University Hospital, Lund, Sweden; 42 Alexandra Balbir-Gurman, Rheumatology Institute Rambam Health Care Campus, Rappaport Faculty Of Medicine, Technion, Haifa, Israel; 49 Raffaele Pellerito, Ospedale Mauriziano, Centro di Reumatologia, Torino, Italy; 50 Luca Idolazzi, Uoc Rheumatology - University Of Verona, Verona, Italy; 52 Christopher Denton, Centre For Rheumatology Royal Free And University College London Medical School, London, United Kingdom; 55 Jelena Colic, Institute of Rheumatology Belgrade, Belgrade, Serbia; 56 Medizinische Universitätsklinik Abt. Ii (Onkologie, Hämatologie, Rheumatologie Immunologie, Pulmonologie, Tübingen, Germany; 57 Katherine Cajiao, Vera Ortiz-SantamariaRheumatology Granollers General Hospital, Granollers Barcelona, Spain; 58 Johannes Pflugfelder, Department of Rheumatology, Marienhospital, Stuttgart, Germany; 59 Dorota Krasowska, Department of Dermatology, Venereology and Pediatric Dermatology, Medical University, Lublin, Poland; 61 Sabine Adler, Kantonsspital Aarau, Dept of Rheumatology and Immunology, Aarau, Switzerland; 68 Tânia Santiago, Department, Centro Hospitalar E Universitário De Coimbra, Coimbra, Portugal; 73 Bojana Stamenkovic, Institute for treatment and rehabilitation Niska Banja, Nis Rheumatology Clinic, Niska Banja, Serbia; 74 Maria De Santis, Irccs Humanitas Research Hospital, Milano, Italy; 78 Lidia P. Ananyeva, V.A. Nasonova Research Institute Of Rheumatology Russian Federation, Mocow, Russia; 81 Ulf Müller-Ladner, Jlu Giessen, Campus Kerckhoff, Department Of Rheumatology And Clinical Immunology, Bad Nauheim, Germany; 86 Merete Engelhart, Department of Rheumatology, University Hospital of Gentofte, Hellerup, Denmark; 87 Gabriela Szücs, University Of Debrecen, Faculty Of Medicine, Department Of Rheumatológy, Debrecen, Hungary; 91 Carlos De La Puente, Servicio De Reumatología, Hospital Ramon Y Cajal Carretera De Colmenar, Madrid, Spain; 93 David Launay, Eric Hachulla, Sébastien Sanges, Univ. Lille, Inserm, CHU Lille, U1286 - INFINITE - Institute for translational Research in Inflammation, F-59000 Lille,France; 96 Andra Balanescu, Department Of Rheumatology - St. Maria Hospital, Carol Davila University Of Medicine And Pharmacy, Bucharest, Romania; 106 Christina Bergmann, Department of Internal Medicine 3, University Hospital Erlangen, Erlangen, Germany; 110 Francesca Ingegnoli, Division Of Rheumatology, Istituto Gaetano Pini Department Of Clinical Sciences & Community Health, University of Milano, Milano, Italy; 112 Luc Mouthon, Department Of Internal Medicine Of Pr Loïc Guillevin Hôpital Cochin, Paris, France; 115 Francesco Paolo Cantatore, Rheumatology Unit - Department of Medical and Surgical Sciences - University of Foggia, Policlinico Ospedali Riuniti di Foggia, Foggia, Italy; 116 Mette Mogensen, University Hospital Of Copenhagen, Department Of Dermatology, Bispebjerg Hospital, Copenhagen, Denmark; 118 Maria Rosa Pozzi, UOSD Reumatologia ASST Monza, Ospedale San Gerardo, Monza, Italy; 120 Piotr Wiland, Department of Rheumatology and Internal Diseases, Wroclaw University of Medicine, Wroclaw, Poland; 122 Marie Vanthuyne, Université Catholique de Louvain, Cliniques Universitaires St-Luc, Brussels, Belgium; 123 Juan Jose Alegre-Sancho, Universitario Dr Peset, Valencia, Spain; 125 Martin Aringer, Division of Rheumatology, University Medical Center Carl Gustav Carus, Dresden, Germany; 126 Ellen De Langhe, University Hospital Leuven, Laboratory Of Tissue Homeostasis And Disease, Department Of Development And Regeneration, Ku Leuven, Leuven, Belgium; 128 Branimir Anic, Division Of Clinical Immunology And Rheumatology, Department Of Internal Medicine, University Of Zagreb, School Of Medicine, University Hospital Center Zagreb, Zagreb, Croatia; 133 Sule Yavuz, Istanbul Bilim University, Dept. Of Rheumatology, Altunizade-Istanbul, Turkey; 135 Carolina De Souza Müller, Hospital De Clinicas Da Universidade Federal Do Parana, Curitiba, Brazil; 137 Svetlana Agachi, Republican Center Of Systemic Sclerosis of Nicolae Testemitanu State University of Medicine and Pharmacy, Sfanta Treime Clinical Hospital, Chisinau, Republic of Moldova; 142 Alberto Cauli, Rheumatology Unit, University Hospital Of Cagliari, Monserrato, Italy; 148 Kamal Solanki, Waikato University Hospital Rheumatology Unit, Hamilton, New Zealand; 152 Esthela Loyo, Departamento Reumatología Hospital Regional Universitario José Ma. Cabral Y Báez Sabana, Santiago, Dominican Republic; 154 Mengtao Li, Department of Rheumatology, Peking Union Medical College Hospital (West Campus), Chinese Academy of Medical Sciences, Beijing, China; 158 Edoardo Rosato, Sapienza University Of Rome-Department Of Translational And Precision Medicine – Centro Di Riferimento Regionale Per La Sclerosi Sistemica, Rome, Italy; 159 Fahrettin Oksel, Ege University Faculty Of Medicine Dept. Internal Medicine Div. Of Rheumatology, Bornova, Izmi, Turkey; 160 Cristina-Mihaela Tanaseanu, Hosp-St. Pantelimon Bucharest, Bucharest, Romania; 161 Rosario Foti, Centre Catania, Uo Reumatologia San Marco Hospital, Catania, Italy; 168 Nihal Fathi, Assiut University Hospital, Assiut University, Rheumatology Department, Assiut, Egypt; 169 Jorge Juan González Martín, Universitario Hm Sanchinarro, Madrid, Spain; 172 Emmanuel Chatelus, University Hospital of Strasbourg-Department of Rheumatology, Hôpital de Hautepierre, Strasbourg, France; 173 Ira Litinsky, Centre Tel-Aviv Sourasky. Rheumatology Institute, Tel-Aviv, Israel; 175 Francesco Del Galdo, Leeds Raynaud’s And Scleroderma Program, Nihr Biomedical Research Centre Leeds Institute Of Rheumatic And Musculoskeletal Medicine, Leeds, United Kingdom; 177 Lesley Ann Saketkoo, New Orleans Scleroderma And Sarcoidosis Patient Care And Research Center, New Orleans, USA; 178 Eduardo Mario Kerzberg, Ramos Mejía Hospital, Buenos Aires, Argentina; 180 Ivan Castellví, Hospital De La Santa Creu I Sant Pau Sant Antoni, Barcelona, Spain; 186 Antonella Marcoccia, Di Riferimento Interdisciplinare Per La Sclerosi Sistemica (Criis), Rome, Italy; 187 Sarah Kahl, Universitätsklinikum Schleswig-Holstein, Campus Lübeck, Innere Medizin/Rheumatologie/Immunologie, Rheumaklinik Bad Bramstedt, Bad Bramstedt, Germany; 188 Vivien M. Hsu, Rutgers-Rwj Scleroderma Program Program Director, New Brunswick, USA; 189 Thierry Martin, Clinical Immunology Internal Medicine. National Referral Center for Systemic Autoimmune Diseases, Nouvel Hopital Civil, Strasbourg, France; 191 Lorinda S Chung, Stanford University School Of Medicine, Stanford, USA; 192 Tim Schmeiser, Krankenhaus St. Josef, Wuppertal-Elberfeld, Germany; 198 Vera Bernardino, De Doencas Autoimunes-Hospital Curry Cabral, Centro Hospitalar Lisboa, Lisboa, Portugal; 199 Gabriela Riemekasten, Klinik für Rheumatologie und Klinische Immunologie, Universitätsklinikum Schleswig-Holstein, Lübeck, Germany; 205 Piercarlo Sarzi Puttini, University Hospital Luigi Sacco, Milano, Italy; 210 Giovanna Cuomo, Uoc Medicina Interna, Università Della Campania, Naples, Italy; 213 Petros Sfikakis, Rheumatology Unit, First Propaedeutic And Internal Medicine, Athens University Medical School, Athens, Greece; 220 Lorenzo Dagna, Unit of Immunology, Rheumatology, Allergy And Rare Diseases, San Raffaele Hospital, Vita-Salute San Raffaele University, Milano, Italy

## Author’s contributions

BM and MK devised the study. ANH and MD worked on the development of data pre-processing workflow. AA developed and implemented the algorithms and models reported in the paper. ANH and AA analyzed and interpreted the data as well as drafted the manuscript. CT contributed to the analysis of feature importance using similarity assessment. BM, MK interpreted the data and supervised and edited the manuscript.

## Acronyms

DLCO: Diffusion Capacity of the Lungs for Carbon Monoxide
FVC: Forced Vital Capacity
SSc: Systemic Sclerosis
ILD: Interstitial Lung Disease
ANP: Attentive Neural Processes
RMSE: Root mean-squared error
NLL: Negative Log-Likelihood
MAE: Mean absolute error

## (A) Additional figures

### B Patient trajectory plots

Figures 12 and 13 demonstrate two patient trajectories with their outcome prediction generated by ANP RNN v2 model. Moreover, each event/visit in the patient’s trajectory (for example Figure 12), is inspected and visualized reporting the top-10 similar (a) continuous, (b) categorical and (c) therapy/medication features between the reference patient and their *closest* (i.e. most *similar*) neighbors averaged for each of these features. We followed the patient similarity approach we described in subsubsection 3.3.1, where for each reference patient representation (latent representation computed by ANP RNN v2 model using past visits up to the prediction event in the timeline), we find the nearest neighbors representation in the training set. These closest neighbors were used to compute an average of the raw input features and compared it to the reference patient’s raw input feature values. In this setup, the raw input features represent a running average of the input features across the past visits in the trajectory up to the event/visit where we predict the future FVC outcome. In a next step we compute the relative distance between the reference patient and nearest neighbors computed average input feature values to identify the top most-similar features. These features are represented in the first three panels in both Figures 12. Additionally, a *k*−NN regression model is fitted on the closest neighbors FVC outcomes to predict the reference patient’s future FVC value. These values are shown in the last panel in the plot where the nearest neighbors FVC values are scattered along with the ANP RNN v2 model prediction, *k*−NN model prediction (representing the average of nearest neighbors FVC values). Moreover, we compute the ratio of unique patients used when selecting nearest neighbor representations. That is, we identify how many distinct patients contributing to the similarity computation, where a ratio of 50/50 means that the 50 timeline representations used in the similarity computation originate from 50 distinct patients. When the numerator is smaller than the denominator, this means at least one patient contributing to multiple timeline representations in the similarity computation. Overall, these plots at the visit level demonstrate the characteristics of the reference patient (i.e. running average of input features up to the prediction event) and their corresponding nearest neighbors. This information helps to offer a link between input features and the predicted outcome shedding some lights on the model’s outcome prediction process. A physician can use these plots to characterize the reference patient and their neighbors status highlighting the raw input features and the potential FVC outcomes (all outcomes and not only the average prediction of the *k*−NN model or the neural model’s prediction).

## C Methods

### C.1 Recurrent neural network (RNN)

We used recurrent neural networks (RNN) that is suited for modeling sequential and temporal data with varying length [28, 29]. RNNs computes a hidden vector at each time step (i.e. state vector *h*_*t*_ at time *t*), representing a history or context summary of the sequence using the input and hidden states vector form the previous time step. This allows the model to learn long-range dependencies where the network is unfolded as many times as the length of the sequence it is modeling. Equation 6 shows the computation of the hidden vector *h*_*t*_ using the input *x*_*t*_ and the previous hidden vector

*h*_*t*−1_ where *ϕ* is a non-linear transformation such as 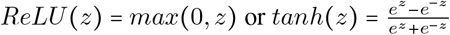.

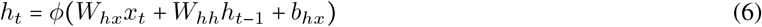

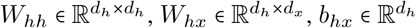, represent the RNN’s weights to be optimized and *d*_*h*_, *d*_*x*_ are the dimensions of *h*_*t*_ and *x*_*t*_ vectors respectively. In this work, we used gated recurrent unit (GRU) [20, 30] to overcome the vanishing/exploding gradient challenges [31, 32, 29] by updating the computation mechanism of the hidden state vector *h*_*t*_ through the specified equations below.

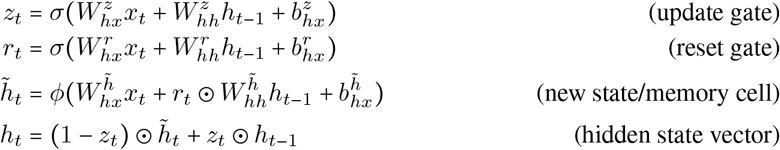

The GRU model computes a reset gate *r*_*t*_ that is used to modulate the effect of the previous hidden state vector *h*_*t*−1_

when computing the new memory vector 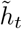. The update gate *z*_*t*_ determines the importance/contribution of the newly generated memory vector 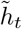 compared to the previous hidden state vector *h*_*t*−1_ when computing the current hidden vector *h*_*t*_ . The weights 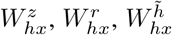 each 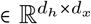 and 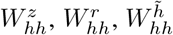 each 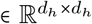 . The biases 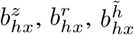 each 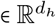 where *d*_*h*_ and *d*_*x*_ are the dimensions of *h*_*t*_ and *x*_*t*_ vectors respectively. The operator *σ* represents the *sigmoid* function, *ϕ* the *tanh* or *ReLU* function, and ⊙ the element-wise product (i.e. Hadamard product). We will refer to the GRU based model by RNN throught the paper.

#### C.1.1 Output Layer

To compute the outcome ŷ_*t*+*δt*_, a fully-connected neural network (i.e. an affine transformation followed by nonlinear function *σ*) is applied to the state vector *h*_*t*_ as in Eq. 7.

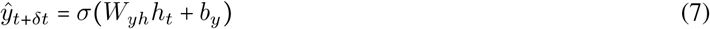

Where 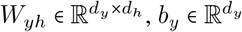.

### C.2 Transformer network

Another model architecture we explored in modelling disease progression is Transformer network [22]. The model has three main blocks: An (1) **Embedding block** that embeds both the *features* and corresponding *absolute position* to a dense vector representation (we also experimented with *time embedding* variation replacing position embedding component). An (2) **Encoder block** that contains (a) a multi-head self-attention layer, (b) layer normalization & residual connections, and (c) feed-forward network. Lastly, an (3) **Output block** representing a regression layer for predicting the subsequent visits FVC value. A formal description of each component of the model is described in their respective sections below.

#### C.2.1 Embedding Block

An embedding matrix *W*_*e*_ is used to map the input *x*_*t*_ to a fixed-length vector representation (Eq. 8)

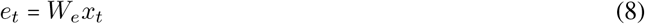

Where 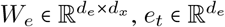, and *d*_*e*_ is the dimension of vector *e*_*t*_.

Similarly, each position *p*_*t*_ in the sequence of visits **x** is represented by 1-of-*T* encoding where *T* is the length of patient’s timeline such that *p*_*t*_ ∈ [0, 1] ^*T*^ . We also experimented with *time embedding* (i.e. binning the visit’s time expressed in years and embedding the corresponding bin) as an alternative to absolute position embedding such that we preserve distances among visits in the sequence. An embedding matrix *W*_*p*_′ is used to map the input *p*_*t*_ to a fixed-length vector representation (Eq. 9)

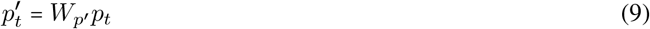

Where 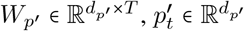 and *d*_*p*_′ is the dimension of vector 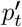 such that *d*_*e*_ and *d*_*p*_′ were equal (denoted by *d* from now on).

Both embeddings *e*_*t*_ and 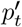 were summed (Eq. 10) to get a unified representation for every element in the sequence (i.e. compute a new sequence *u* = [*u*_1_, *u*_2_, ⋯, *u*_*T*_] represented by matrix *U* ∈ ℝ^*T* ×*d*^, where *u*_*t*_ ∈ℝ^*d*^, ∀*t* ∈[1, ⋯, *T*]).

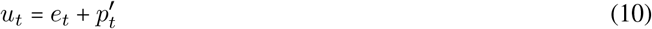

#### C.2.2 Encoder Block: Multihead Self-Attention Layer with Causal Mask

We used a multi-head self-attention approach where multiple single-head self-attention layers are used in parallel (i.e. simultaneously) to process each input vector *u*_*t*_. The outputs from every single-head layer are concatenated and transformed to generate a fixed-length vector using an affine transformation. The single-head self-attention approach *SHA* [22] performs linear transformation to the input vectors using three separate matrices: (1) a queries matrix 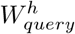, (2) keys matrix 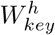, and (3) values matrix 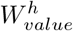. The input matrix *U* is mapped using these matrices to

compute three new matrices (Eq. 11, 12, and 13)

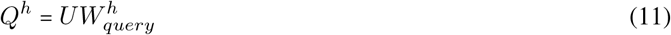

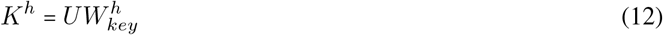

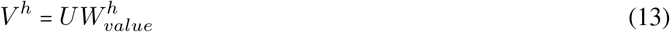

where 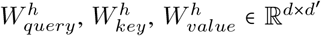, *Q*^*h*^, *K*^*h*^, *V* ^*h*^ ∈ ℝ^*T* ×*d*^′ are query, key and value matrices, *d*^′^ is the common embedding dimension, and h is indexing attention heads in *H* multi-head setting. In a second step, attention scores are computed using the pairwise similarity between the query and key vectors for each position *t* in the sequence. The similarity is defined by computing a scaled dot-product between the pairwise vectors. A *CausalMask* (16) is used to restrict information access to the past visits only offering a *causal* attention layer. This is done by element-wise multiplying ⊙ the unnormalized similarity matrix with a matrix composed of 1s on the lower triangular part and −∞ on the upper triangular part. After *softmax* operation the attention scores will form a normalized lower triangular matrix that is used to perform a weighted sum with the value vectors (Eq. 15) to generate a new matrix representation.

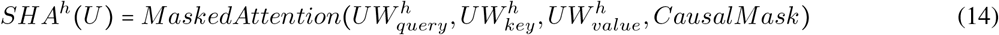

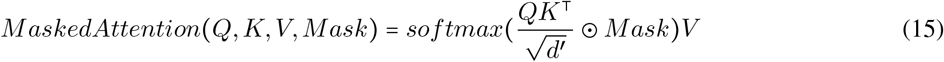

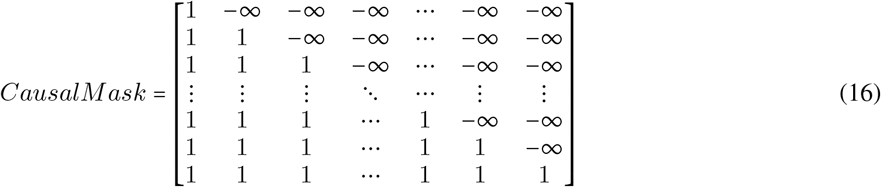

In a multi-head setting with *H* number of heads, multiple *SHA* transformations are applied separately to be later concatenated (⊕) along features dimension and then transformed using affine transformation (Eq. 17) such that *W*_*unify*_ ∈ ℝ^*d*^′_*H*×*d*_ and *b*_*unify*_ ∈ ℝ^*d*^.

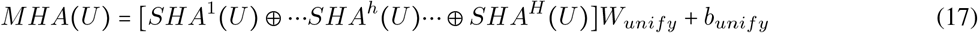

#### C.2.3 Encoder Block: Layer Normalization & Residual Connections

Residual/skip connections [42] and layer normalization [43] are used during training between two sub-layers: multihead attention and the feed-forward layer. This is to improve the gradient flow in layers and to ameliorate the “covariate-shift” problem by re-standardizing the computed vector representations. *LayerNorm* function will standardize the input vector using the mean *μ*_*t*_ and variance 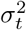 along the features dimension *d* of an input vector *u*_*t*_ and apply a scaling *γ* and shifting step *β* (Eq. 20). *γ* and *β* are learnable parameters and *ϵ* is small number added for numerical stability. Hence, new output ũ_*t*_ is computed using Eq. 21 to generate a new matrix *Ũ* ∈ ℝ^*T* ×*d*^.

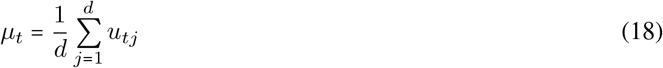

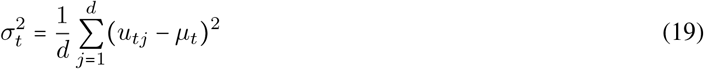

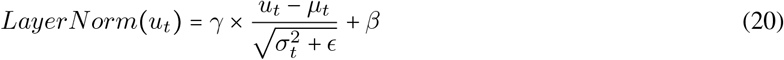

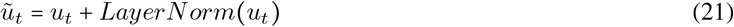

#### C.2.4 Encoder Block: FeedForward Layer

The final sub-layer in encoder block is a feed-forward network consisting of two affine transformation matrices and non-linear activation function is used to further compute/embed the learned vector representations from previous layers (i.e. *Ũ* = [ũ ; ũ ; ⋯; ũ] ∈ℝ^*T* ×*d*^). The first transformation (Eq. 22) uses *W*_*MLP* 1_ ∈ℝ^*ξd*^′_×*d*_ and *b*_*MLP* 1_ ∈ ℝ^*ξd*^′ to transform ũ_*t*_ to new vector ∈ ℝ^*ξd*^ where *ξ* ∈ ℕ is multiplicative factor. A non-linear function such as *ReLU* is applied followed by another affine transformation using *W*_*MLP* 2_ ∈ ℝ^*d*×*ξd*^′ and *b* _*MLP* 2_ ∈ℝ^*d*^ to obtain vector *f*_*t*_ ∈ℝ^*d*^. A layer normalization plus residual connection (Eq. 23) is applied to obtain 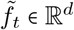 and consequently matrix 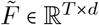.

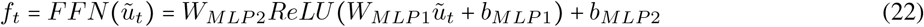

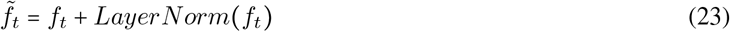

At this point, the *encoder* block operations are done and multiple encoder blocks can be stacked in series for *E* number of times. In our experiments, *E* was a hyperparameter that was empirically determined using a validation set (as the case of the number of attention heads *H* used in self-attention layer).

#### C.2.5 Output Layer

To compute the outcome ŷ_*t*+*δt*_, a fully-connected neural network is applied to 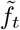 Eq. 24.

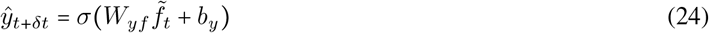

Where 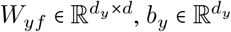.

### C.3 Attentive Neural Processes (ANP)

Attentive Neural Processes (ANP) [19] is an extension to Neural Processes [33] an approach that learns a distribution over functions mapping the input to output from a training set (i.e. learning a posterior distribution over *f* the underlying function mapping input to output) that is further used to make inference for test points. ANP defines an infinite family of conditional distributions conditioning on arbitrary number of *contexts* (i.e. set of input-output pairs) (**x**_*C*_, **y**_*C*_) = {(*x*_1_, *y*_1_), (*x*_2_, *y*_2_), ⋯, (*x*_*C*_, *y*_*C*_)} to model arbitrary number of *targets* (**x**_*M*_, **y**_*M*_) = {(*x*_1_, *y*_1_), ( *x*_2_, *y*_2_), ⋯,(*x*_*C*_, *y*_*C*_), ⋯,(*x*_*M*_, *y*_*M*_) } invariant to the ordering of both the contexts and targets where *C* ⊂ *M* Eq. 25. In this work, we adapt ANP to model patient trajectories (i.e. timeseries data) where causal temporal ordering is preserved and we describe the adaptation of the modeling approach from this perspective.

ANP comprises of an (1) **Encoder block** that uses two paths (a) *deterministic* and (b) *latent path*, and (2) **Decoder block** that maps the computed representation from the encoder block to the the target output (Figure 14).

**Figure 14:**
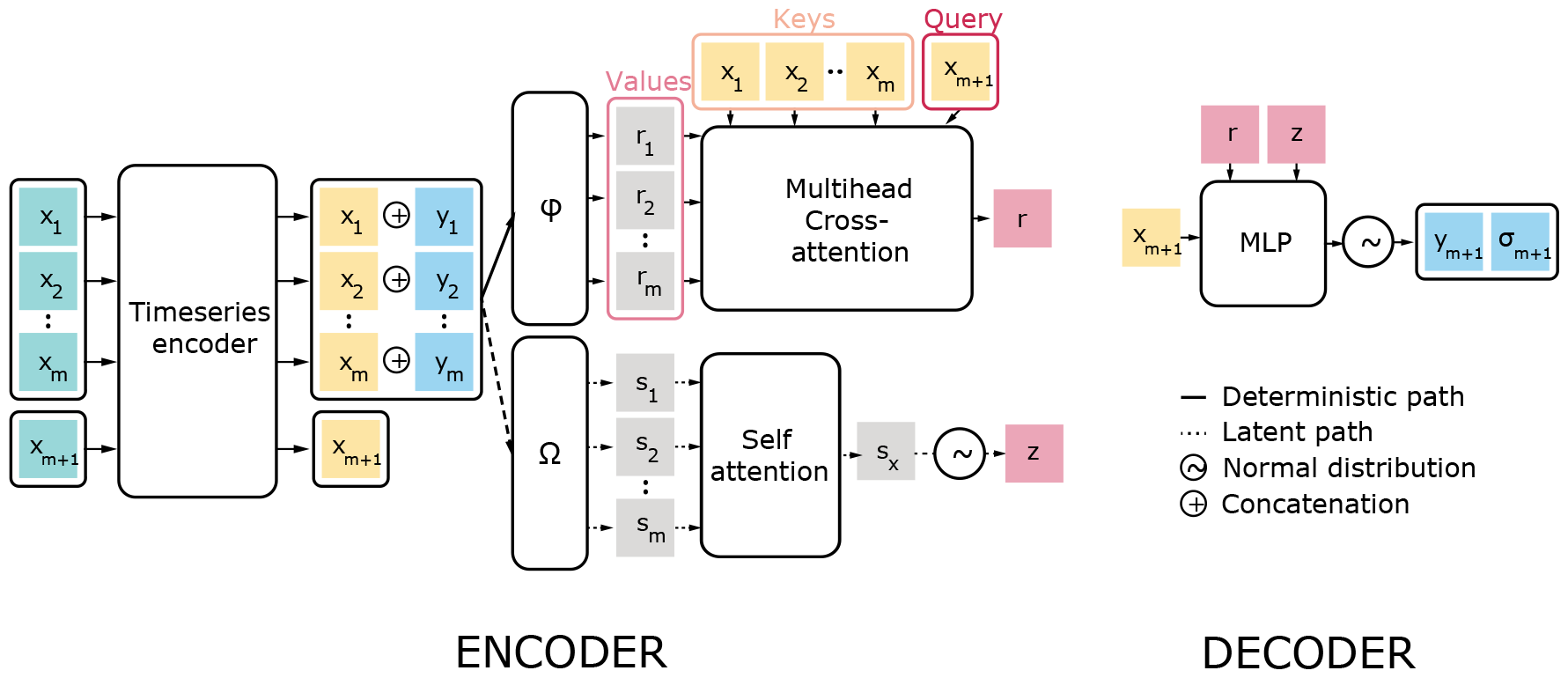
Attentive Neural Processes architecture for timeseries

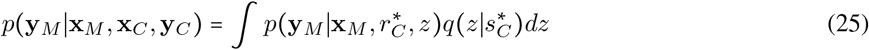

#### C.3.1 Encoder: Deterministic path

The deterministic path uses a deterministic function (i.e. encoder Φ **x**_*C*_, **y**_*C*_) that takes *C* input-output context visits (**x**_*C*_, **y**_*C*_) to generate **r**_*C*_ ∈*ℝ*^*C*×*d*^ representations Eq. 26. A cross-attention layer similar to the single head attention layer (Eq.28) takes a set of targets **x**_*M*_ as queries to attend to set of contexts **x**_*C*_ (as keys) in order to obtain attention scores (normalized similarity matrix) that is further used to weight the **r**_*C*_ vectors (acting as values) to generate a fixed-length summary vector 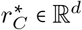 capturing the local structure for the query-specific representation (Eq. 27).

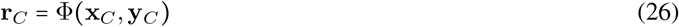

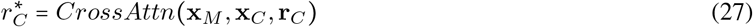

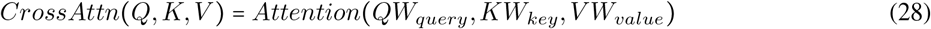

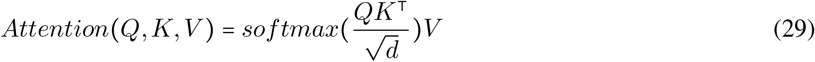

#### C.3.2 Encoder: Latent path

The latent path is used to compute a global latent representation *z* to account for the uncertainty in the output prediction of targets **y**_*M*_ given the contexts (**x**_*C*_, **y**_*C*_) . *z* would capture the global structure by modelling the different realizations of the underlying stochastic process generating the data, and providing a latent summary complementing the deterministic summary representation 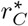 . This is done by first passing the contexts (**x**_*C*_, **y**_*C*_) to an encoder Ω to compute **s**_*C*_ vectors ∈ ℝ^*C*×*d*^ (Eq. 30). These vectors are aggregated using mean pooling or attention layer that uses a learnable query vector *q*, and **s**_*C*_ vectors as key-value pairs. Then an affine transformation followed by nonlinear activation *κ*, is applied to generate 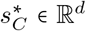(Eq. 31). The mean and variance vectors *μ*_*z*_ (Eq. 32) and *σ*_*z*_ (Eq. 33) are computed using two separate affine transformations 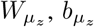 and 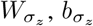 respectively using 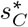 vector to parameterize a factorized Gaussian 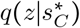 Eq. 34. *ξ* is hyperparameter ∈ [0., −1]. used to bound the variance.

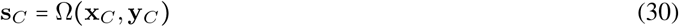

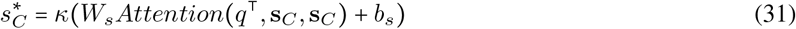

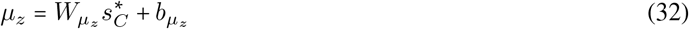

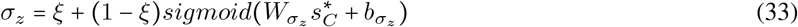

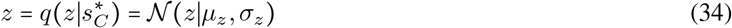

#### C.3.3 Decoder block

The computed deterministic 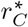 and latent 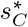 vector representations from the contexts in addition to targets **x**_*M*_ are concatenated and embedded (Eq. 35) using an affine transformation *W*_*v*_ ∈ *ℝ*^3*d*×*d*^ and *b*_*v*_ ∈ℝ^*d*^. A feed-forward layer with layer normalization and residual connections similar to the encoder layer in (section C.2.4) is used and the output is passed to two separate affine transformations to generate two vectors *μ*_*y*_ and *σ*_*y*_ representing the mean and variance parametrizing a factroized Gaussian distributions of the outcomes across the targets **x**_*M*_ .

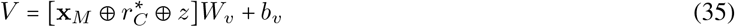

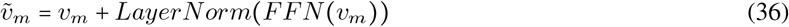

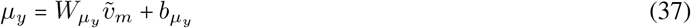

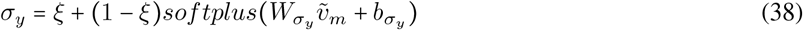

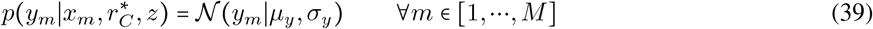

In this study, we first used a time series encoder that embeds the raw input for both the context and target events, then pass the learned representations to the encoder blocks Φ and Ω. We experimented with two model variations: the first used gated RNN for all the encoder blocks and is denoted by ANP RNN, and the second used transformer based encoder blocks and is denoted by ANP transformer.

## D Models’ hyperparameter options

**Table 7:**
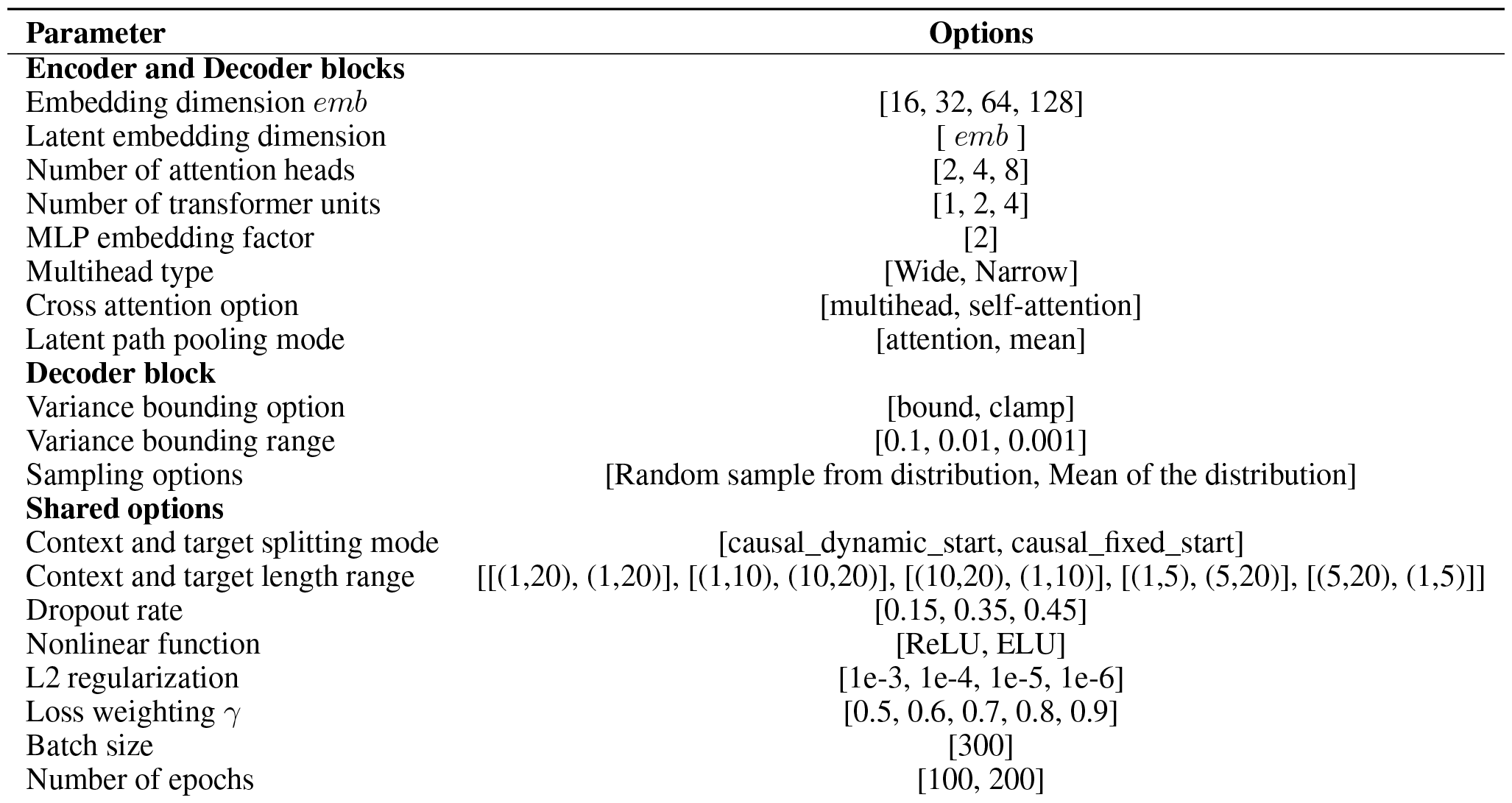
Hyperparameter space of attentive neural process with transformer blocks.

| Parameter | Options |
| --- | --- |
| **Encoder and Decoder blocks** |  |
| Embedding dimension $emb$ | [16, 32, 64, 128] |
| Latent embedding dimension | [ $emb$ ] |
| Number of attention heads | [2, 4, 8] |
| Number of transformer units | [1, 2, 4] |
| MLP embedding factor | [2] |
| Multihead type | [Wide, Narrow] |
| Cross attention option | [multihead, self-attention] |
| Latent path pooling mode | [attention, mean] |
| **Decoder block** |  |
| Variance bounding option | [bound, clamp] |
| Variance bounding range | [0.1, 0.01, 0.001] |
| Sampling options | [Random sample from distribution, Mean of the distribution] |
| **Shared options** |  |
| Context and target splitting mode | [causal_dynamic_start, causal_fixed_start] |
| Context and target length range | [[ (1,20), (1,20) ], [ (1,10), (10,20) ], [ (10,20), (1,10) ], [ (1,5), (5,20) ], [ (5,20), (1,5) ]] |
| Dropout rate | [0.15, 0.35, 0.45] |
| Nonlinear function | [ReLU, ELU] |
| L2 regularization | [1e-3, 1e-4, 1e-5, 1e-6] |
| Loss weighting $\gamma$ | [0.5, 0.6, 0.7, 0.8, 0.9] |
| Batch size | [300] |
| Number of epochs | [100, 200] |

**Table 8:**
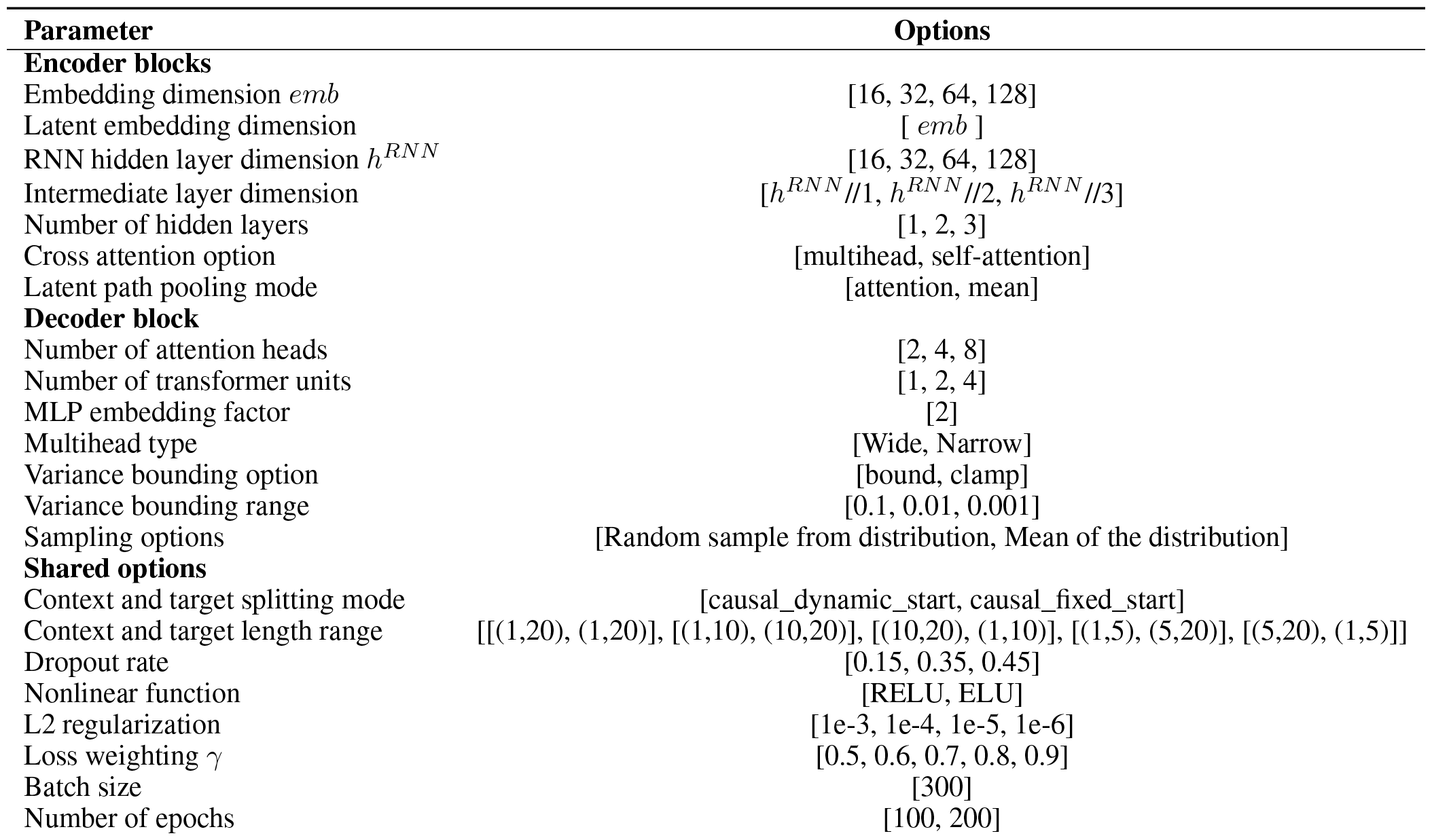
Hyperparameter space of attentive neural process with RNN blocks.

**Table 9:** Hyperparameter space of RNN model.

| Parameter | Options |
| --- | --- |
| Embedding dimension | [16, 32, 64, 128] |
| RNN hidden layer dimension $h^{RNN}$ | [16, 32, 64, 128] |
| Intermediate layer dimension | $[h^{RNN} // 1, h^{RNN} // 2, h^{RNN} // 3]$ |
| Number of hidden layers | [1, 2, 3] |
| Dropout rate | [0.15, 0.35, 0.45] |
| RNN cell type | [GRU, LSTM] |
| Nonlinear function | [ReLU, ELU] |
| L2 regularization | [1e-3, 1e-4, 1e-5, 1e-6] |
| Batch size | [300] |
| Number of epochs | [100, 200] |

**Table 10:** Hyperparameter space of transformer model.

| Parameter | Options |
| --- | --- |
| Embedding dimension $emb$ | [16, 32, 64, 128] |
| Number of attention heads | [2, 4, 8] |
| Number of transformer units | [1, 2, 4] |
| Dropout rate | [0.15, 0.35, 0.45] |
| Nonlinear function | [ReLU, ELU] |
| MLP embedding factor | [2] |
| Multihead type | [Wide, Narrow] |
| Batch size | [300] |
| Number of epochs | [100, 200] |

**Table 11:** Hyperparameter space of RandomForestRegressor, HistGradientBoostingRegressor, ^*^parameter only for the HistGradientBoostingRegressor.

| Parameter | Options |
| --- | --- |
| Number of estimators | list(range(10,501,30)) |
| Maximum number of features | ['sqrt'] |
| Maximum depth | [2, 10, 100, not restricted] |
| Minimum sample split | [2, 5, 10] |
| Minimum samples required at each leaf node | [1, 5, 10, 20] |
| L2 regularization* | [1e4, 1e3, 1e2, 1e1, 1., 1e-1, 1e-2, 1e-3, 1e-4, 1e-6] |

**Table 12:** Hyperparameter space of XGBoost.

| Parameter | Options |
| --- | --- |
| Minimum child weight | [1, 2, 4, 8, 10, 16] |
| Gamma | [0, 1, 2, 4, 6, 8] |
| Subsample ratio of the training instances | [0.5, 1.0] |
| Subsample ratio of columns | [0.5, 1.0] |
| Maximum depth of a tree | [6, 9, 15] |
| Learning rate | [0.05, 0.1, 0.2, 0.3, 0.5] |
| Number of estimators | list(range(10, 501, 30)) |
| Lambda | [1e4, 1e3, 1e2, 1e1, 1., 1e-1, 1e-2, 1e-3, 1e-4, 1e-8] |
| Alpha | [1e4, 1e3, 1e2, 1e1, 1., 1e-1, 1e-2, 1e-3, 1e-4, 1e-8] |

**Table 13:** Hyperparameter space of Ridge, Lasso, ElasticNet, ^*^parameter only for the ElasticNet.

| Parameter | Options |
| --- | --- |
| Alpha | [1e4, 1e3, 1e2, 1e1, 1., 1e-1, 1e-2, 1e-3, 1e-4, 1e-6] |
| L1 ratio* | numpy.arange(0, 1.1, 0.1) |

## E Features

**Table 14:**
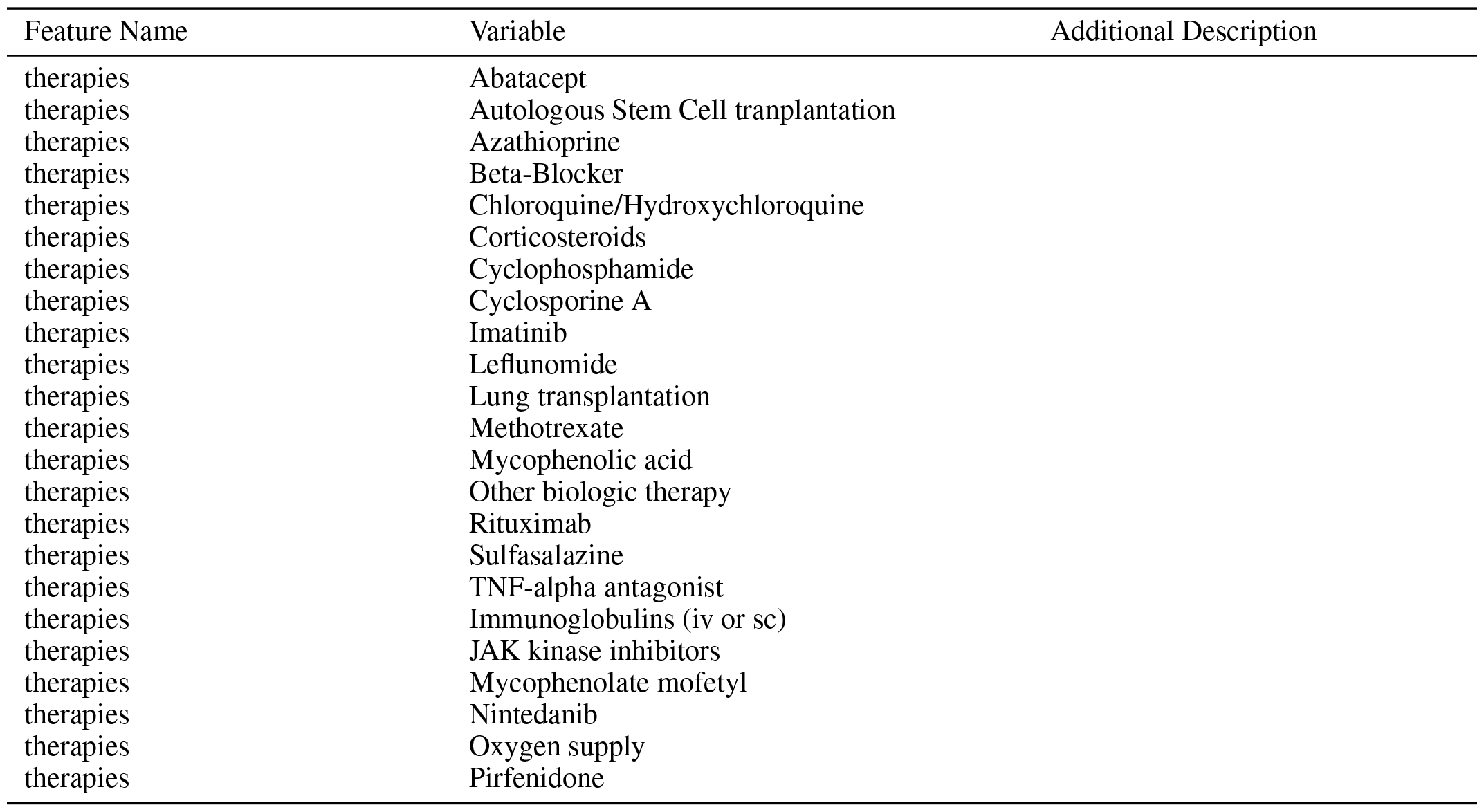
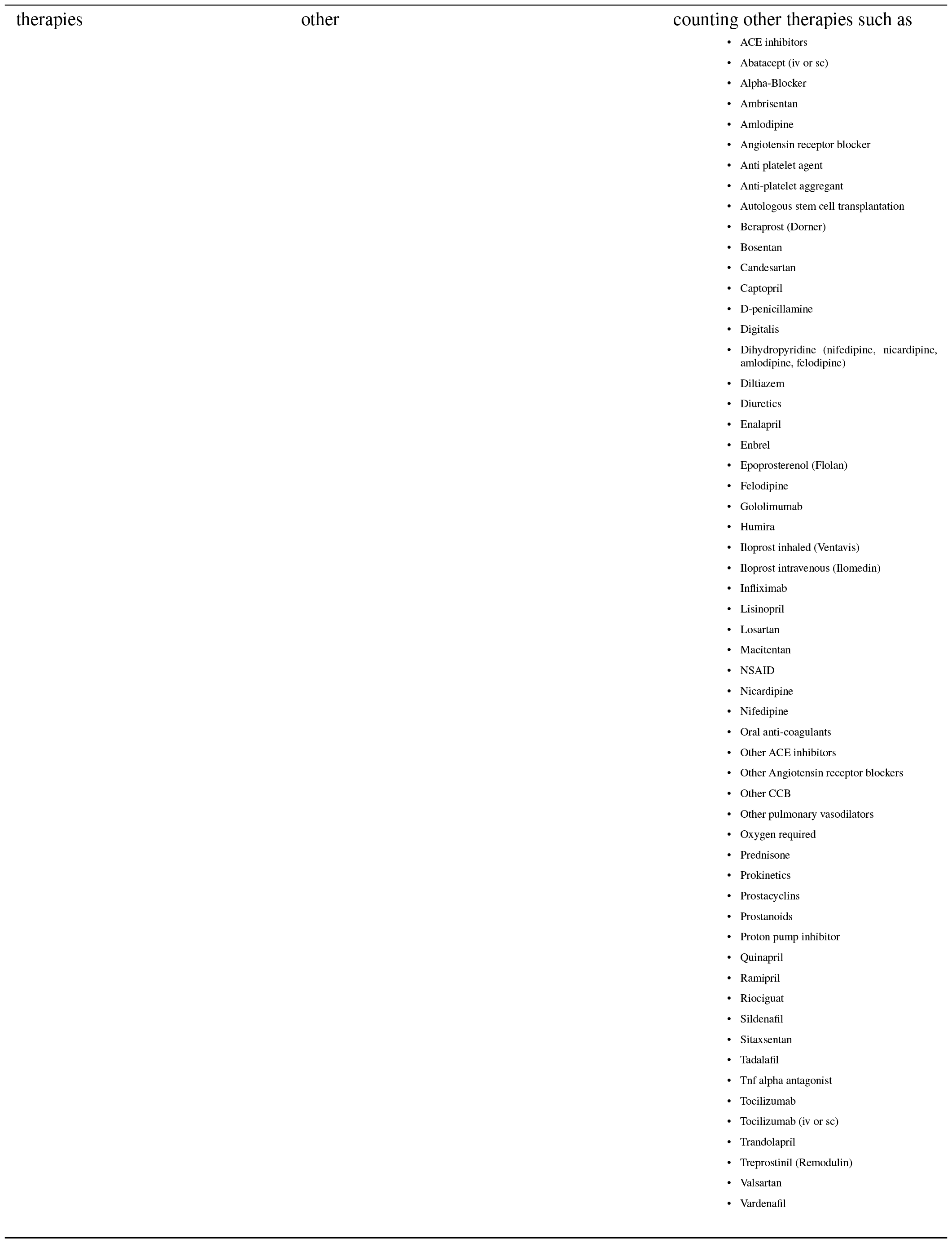
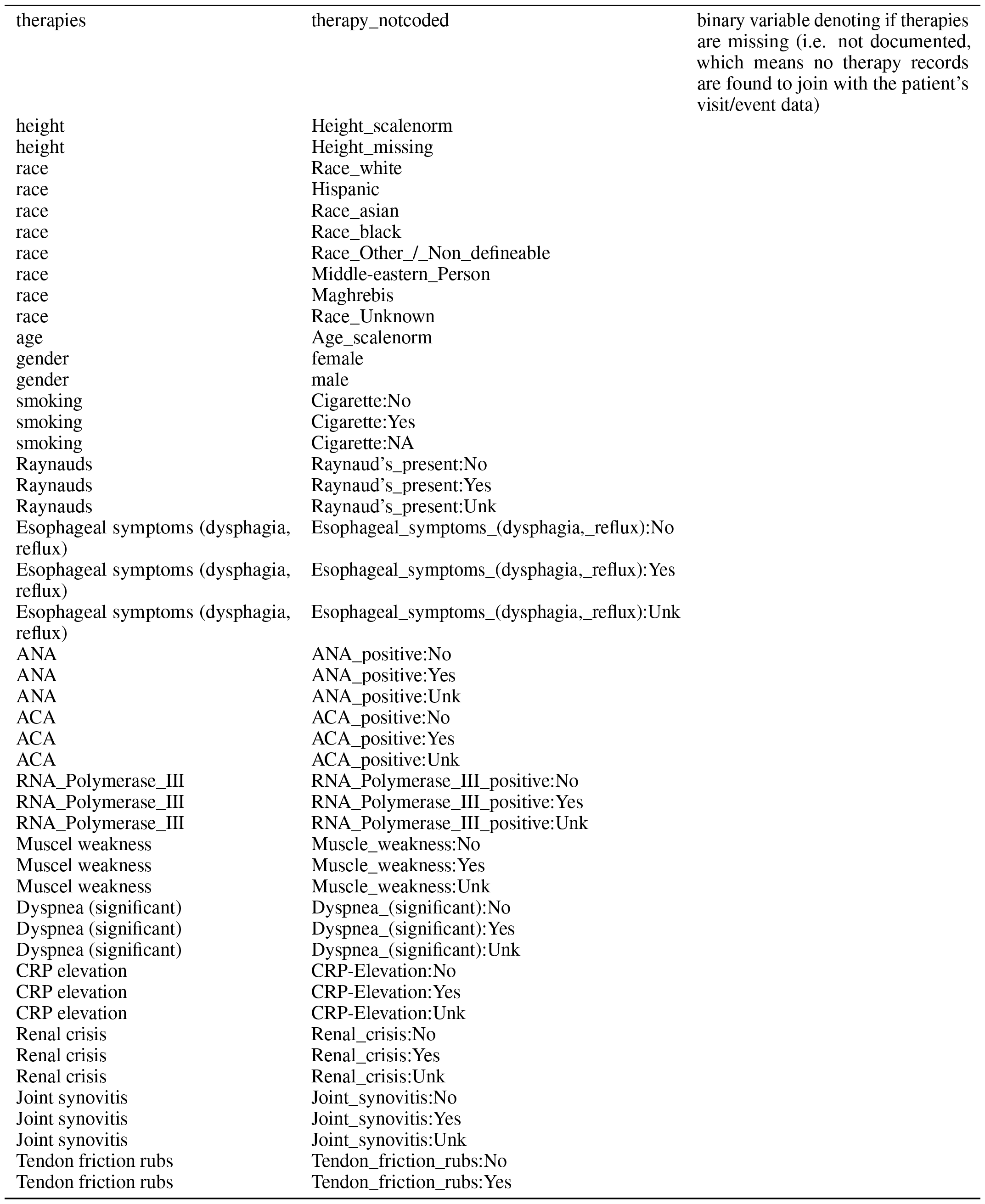
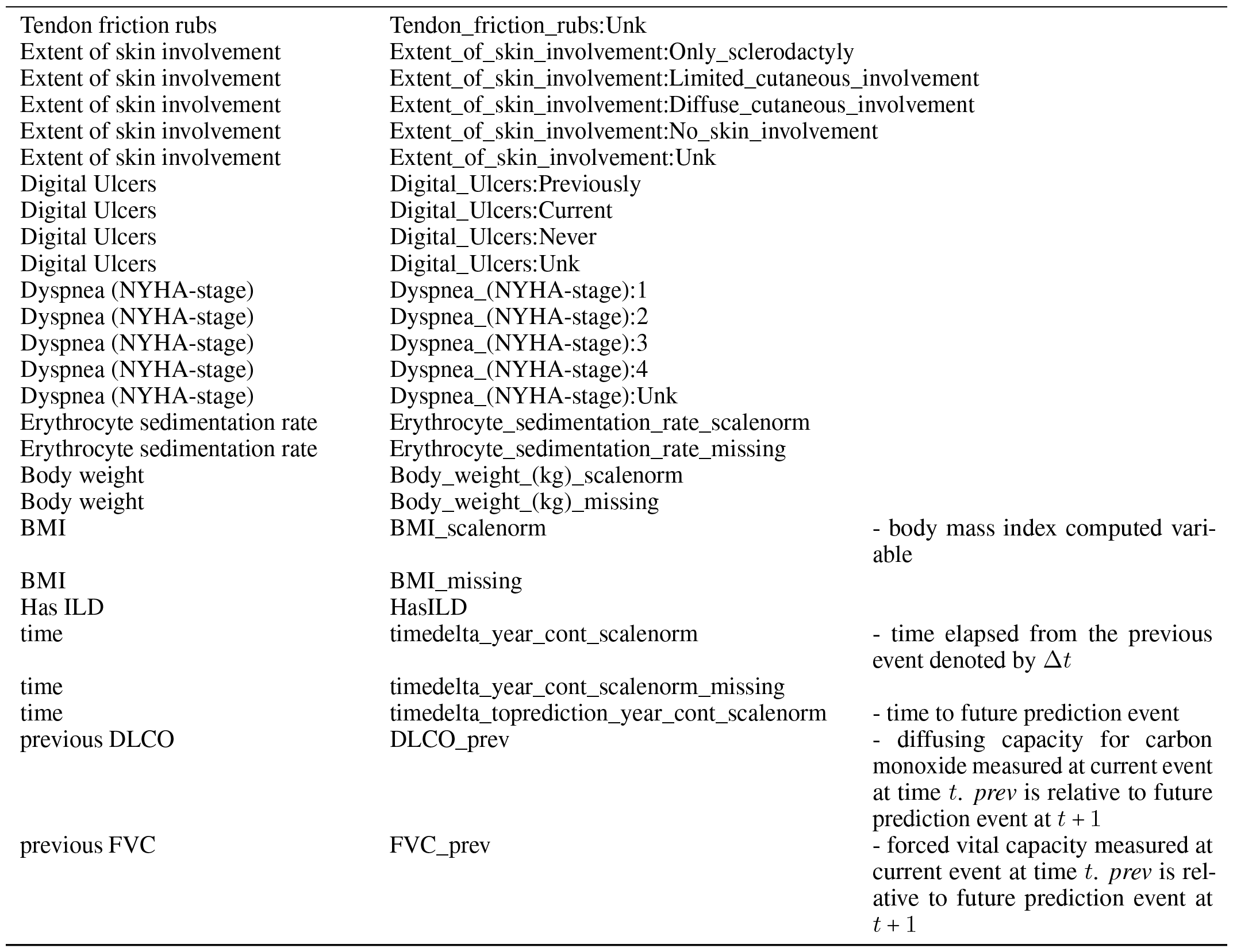
Features/variables used in neural models.

